# Quantification of *Legionella pneumophila* in building potable water systems: a meta-analysis comparing qPCR and culture-based detection methods

**DOI:** 10.1101/2024.05.02.24306716

**Authors:** Émile Sylvestre, William J. Rhoads, Timothy R. Julian, Frederik Hammes

**Affiliations:** Eawag, Swiss Federal Institute of Aquatic Science and Technology, Dübendorf, CH-8600, Switzerland; Sanitary Engineering, Delft University of Technology, Stevinweg 1, 2628 CN, Delft, the Netherlands; Black & Veatch, 8400 Ward Parkway, Kansas City, MO 64114; Swiss Tropical and Public Health Institute, CH-4123 Allschwil, Switzerland; University of Basel, CH-4055 Basel, Switzerland

**Keywords:** *Legionella pneumophila*, quantitative PCR, culture-based method, building potable water systems, quantitative microbial risk assessment (QMRA)

## Abstract

Quantitative polymerase chain reaction (qPCR) offers a rapid, automated, and potentially on-site method for quantifying *L. pneumophila* in building potable water systems, complementing and potentially replacing traditional culture-based techniques. However, the application of qPCR in assessing human health risks is complicated by its tendency to overestimate such risks due to the detection of genomic copies that do not correspond to viable, infectious bacteria. This study examines the relationship between *L. pneumophila* measurements obtained via qPCR and culture-based methods, aiming to understand and establish qPCR-to-culture concentration ratios needed to inform associated health risks. We developed a Poisson lognormal ratio model and a random-effects meta-analysis to analyze variations in qPCR-to-culture ratios within and across sites. Our findings indicate these ratios typically vary from 1:1 to 100:1, with ratios close to 1:1 predicted at all sites. Consequently, adopting a default 1:1 conversion factor appears necessary as a cautious approach to convert qPCR concentrations to culturable concentrations for use in models of associated health risks, for example, through quantitative microbial risk assessment (QMRA) frameworks. Where this approach may be too conservative, targeted sampling and the applications of viability-qPCR could improve the accuracy of qPCR-based QMRA. Standardizing qPCR and culture-based methods and reporting site-specific environmental factors that affect the culturability of *L. pneumophila* would improve the understanding of the relationship between the two methods. The ratio model introduced here shifts us beyond simple correlation analyses, facilitating investigations of temporal and spatial heterogeneities in the relationship. This analysis is a step forward in the integration of QMRA and molecular biology, as the framework demonstrated here for *L. pneumphila* is applicable to other pathogens monitored in the environment.

## 1 Introduction

*Legionella pneumophila* is widely regarded as the primary causative agent of Legionnaires’ disease (Phin et al., 2014), and therefore, accurately quantifying *L. pneumophila* concentrations in water is central to prevention strategies. Culture-based methods defined by standards like ISO 11731:2017, NF T90-431:2017, and ASTM D8429-21 are commonly used to quantify *L. pneumophila* concentrations in colony-forming units (CFU) or most probable number (MPN). Such cultivation-derived data then serve as inputs for quantitative microbial risk assessment (QMRA), enabling predictions of probabilities of infection upon exposure (Armstrong and Haas, 2007; Armstrong and Haas, 2008). However, the widespread application of culture-based methods is hindered by several limitations (National Academies of Sciences and Medicine, 2020), including their time-consuming nature (8–14 days to obtain results), low processing throughput, the need for specialized cultivation expertise, and concerns about so-called viable but not culturable (VBNC) bacteria.

In some circumstances, molecular techniques like quantitative polymerase chain reaction (qPCR) present a solution to some of these limitations. With their capacity for high-throughput, rapid, and specific *L. pneumophila* genome copies (GC) quantification, as well as benefits such as automation and on-site implementation, qPCR methods offer a compelling tool to use in conjunction with cultivation (Ahmed et al., 2019; Simard and Doyer, 2022). While standardised qPCR is well established for *L. pneumophila* detection (AFNOR, 2010; Anonymous, 2012), cultivation will most likely remain the dominant reference method for the foreseeable future. Hence, a comprehensive understanding of the relationship between cultivation and qPCR data is crucial for advancing the use of qPCR for monitoring and risk assessment.

Studies comparing *L. pneumophila* concentrations obtained through qPCR and culture-based methods within building water systems often report correlations (Collins et al., 2017; Joly et al., 2006). However, the relationship between qPCR and culture is not expected to be always conserved, both between and within sites, due to various factors, such as environmental conditions affecting the viability and cultivability of the organism (e.g., disinfectant concentration, water temperature, age of pipes/biofilms, water flow conditions) (Delgado-Viscogliosi et al., 2009; Donohue, 2021; Grimard-Conea et al., 2022), and method-specific performances related to analytical recovery and detection/quantification limits (Lee et al., 2011). In some instances, qPCR might overestimate human health risks by capturing all DNA, including DNA from all cells (culturable, VBNC, and dead) and also free DNA. Emerging techniques, which target DNA within intact cells, often referred to as viability-qPCR, may better align concentration estimates with associated health risks (Nisar et al., 2023) but are still not widely adopted in qPCR-based applications. Additionally, bacterial aggregation in environmental samples (Haas and Heller, 1986; Haas and Heller, 1988) may result in qPCR yielding higher concentrations than cultivation. Thus, employing a ratio as a model could help identify situations with significant discrepancies between the two quantification methods (Lee et al., 2011), improving our understanding of their interplay.

This study presents a systematic review and meta-analysis of the qPCR:culture concentration ratio for *L. pneumophila* in building potable water systems. Our objectives were to (i) develop statistical models to describe the variability in the qPCR:culture relationship both within and across various studies to generate a comprehensive understanding of literature data (ii) to apply these models in interpreting qPCR results within a QMRA framework, and (iii) to evaluate and interpret the factors influencing the qPCR:culture ratio.

## 2 Systematic review

The scientific studies included in this work were identified, screened, and selected following the guidelines established in the Preferred Reporting Items for Systematic Reviews and Meta-Analysis (PRISMA) framework (Page et al., 2021). A systematic review protocol was initially developed and amended during the study (Supplementary Material, S1).

### 2.1 Eligibility criteria

This review was limited to the results obtained from the analysis of water samples collected in full-scale building potable water systems. Eligible studies necessarily collected quantitative data on the concentrations of *L. pneumophila* in water using both molecular methods (qPCR, viability-qPCR) and culture-based methods (ISO 11731, Legiolert) within paired water samples, as the outcome of interest is the qPCR:culture concentration ratio. Both most probable number (MPN) and colony forming units (CFU) were included for the quantification of cultivable concentrations because both approaches aim to assess viable *L. pneumophila* populations. This inclusion allows for a more comprehensive analysis across a broader range of studies.

Only studies including data sets with at least 20% of quantifiable samples were selected to ensure reliable statistical assessments. Studies reporting building-specific data and pooled data from multiple buildings were included in the review, but only building-specific data were used for meta-analysis. Studies using alternative molecular methods, such as digital droplet PCR (ddPCR) and reverse-transcription PCR (RT-qPCR), were excluded. The search was restricted to peer-reviewed journals and governmental reports published in English, French, and German.

### 2.2 Search strategy

Backward and forward searches were conducted by the first and second reviewers (hereafter referred to as WR and ES, respectively), starting with a narrative review paper on methods comparison for *Legionella* enumeration (Whiley and Taylor, 2014) and the National Academies of Science, Engineering and Medicine (NASEM) report Management of *Legionella* in Water Systems (National Academies of Sciences and Medicine, 2020). Based on the information gathered from these sources, an electronic search was conducted on PubMed, Scopus, and Web of Science using the following search string:

“(Legio* AND qPCR AND culture) OR (Legio* AND viability PCR AND culture) OR (Legio* AND EMA AND culture) OR (Legio* AND PMA AND culture) OR (Legio* AND Methods AND Compare).”

Title and abstract screenings were conducted by WR, with verification of excluded articles by ES. WR and ES duplicated the full-text screening, and discrepancies were resolved through discussion. Reasons for excluding studies were recorded. A collaborative spreadsheet software (Google Sheets, Alphabet Inc) was used to retrieve citations, screen citations, and record reasons for study exclusion.

### 2.3 Data extraction

Water system information, sample processing details, and measurement information, as shown in Table 1, were extracted from the selected studies by WR, ES, and a research intern. Additionally, to evaluate the quality control and assurance of qPCR analyses, we verified whether studies reported methods for generating standard curves for qPCR, control measures for nucleic acid extraction, PCR detection, and inhibition. For cultivation methods, we verified whether standardized protocols were applied. The extracted data were verified by a second reviewer (WR or ES) in all studies to ensure accuracy. For eight studies, concentrations or counts were not provided or reported in tables. For these studies, WebPlotDigitizer, a data extraction program with nearly perfect accuracy (Aydin and Yassikaya, 2022), was used in manual mode to extract approximated concentrations from relevant figures by magnifying them at 500%. All data extracted from studies were stored in the collaborative spreadsheet software, facilitating collaborative document editing and ensuring consistency.

**Table 1.** Reporting elements necessary for evaluating the relationship between concentrations of *Legionella pneumophila* measured by qPCR and culture-based methods in building potable water systems.

| Category | Parameter |
| --- | --- |
| Water system information | Building type<br>Water system (e.g., hot/cold)<br>Outlet<br>Draw (e.g., first draw, flushed) |
| Water treatment | Preventive control measure(s)<br>Curative control measure(s) |
| Water quality parameter | Total chlorine<br>Free chlorine<br>Monochloramine<br>Water temperature |
| Sample processing information | Sampling location and date<br>Sample type<br>Sample volume for qPCR<br>Sample volume for cultivation<br>Sample dilution/concentration factor for qPCR<br>Sample dilution/concentration factor for cultivation<br>Volume processed for qPCR<br>Volume processed for culture |
| Measurement information<sup>a</sup> | qPCR method<br>Gene target for qPCR<br>Number of gene copies within the genome<br>Specificity of the qPCR assay<br>Culture-based method (e.g., CFU, MPN)<br>Count of *Legionella pneumophila* per processed volume for qPCR<sup>b</sup><br>Count of *Legionella pneumophila* per processed volume for cultivation<sup>b</sup><br>Limits of detection (LOD) for qPCR<br>Limits of detection (LOD) for cultivation<br>Limit of quantification (LOQ) for qPCR<br>Limit of quantification (LOQ) for cultivation<br>Recovery rates for qPCR (whole process, nucleic acid extraction, PCR detection)<br>Recovery rates for cultivation |

<sup>a</sup>This list does not include all quality control and assurance elements necessary for qPCR and cultivation analyses of environmental samples. <sup>b</sup>When counts were unavailable, reported concentrations were extracted.

## 3 Statistical analyses

### 3.1 Variability of the qPCR:culture ratio within a building

To estimate how much the ratio between the results of the two methods (qPCR and cultivation) varies within each building, we adopted the statistical method previously developed by Sylvestre et al. (2021). This method involves comparing two sets of results that follow a specific probability distribution, known as a Poisson−lognormal distribution. This model assumes that counts *x* (CFU for culture and GC for qPCR) are randomly (Poisson) distributed in each water sample of volume *V* and concentration *c*. The probability of finding *x* organisms in the water sample is thus:

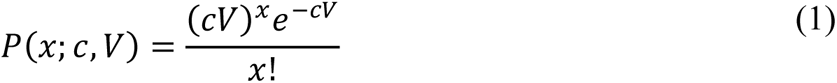

Organisms have a concentration *c*, and their expected number in a sample volume *V* is *cV*. The concentration *c* is likely to vary in time or space. For MPN methods, Eq. 1 was used as an approximation method because primary studies did not report positive and negative well counts. When such data are reported, the MPN method uncertainty can be modelled more accurately using the binomial and Poisson distributions (Haas et al., 2014).

This variation in concentration *c* can be described by a lognormal distribution, which has the following probability density function:

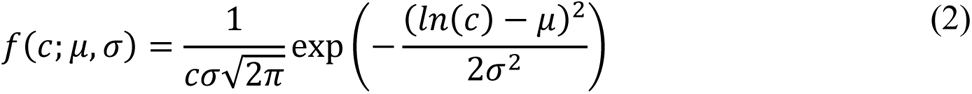

where *μ* and *σ* are, respectively, the mean and the standard deviation of the underlying normal distribution on the logarithmic scale. If Eq. 2 describes the variation of *c* in Eq. 1, then the count, *x*, follows a Poisson−lognormal distribution. The goodness of fit of the Poisson−lognormal distribution was compared to the goodness of fit of an alternative model, the Poisson−gamma distribution, using the marginal deviance information criterion (mDIC) (Quintero and Lesaffre, 2018). Results indicated that the lognormal distribution better described concentration variations for qPCR data (Table S1) and cultivation data (Table S2); therefore, the Poisson lognormal distribution was chosen for modelling *L. pneumophila* concentrations and their ratios. As the *mip* gene, which is used to quantify *L. pneumophila* via qPCR, exists in a single copy within the genome of *L. pneumophila* (Engleberg et al., 1989), methods can be compared by directly taking the ratios of CFUs to gene copies.

The distribution of the ratio of two lognormal random variables is also lognormally distributed with a mean of:

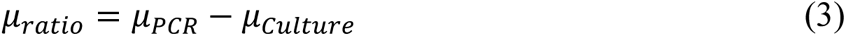

and a variance of:

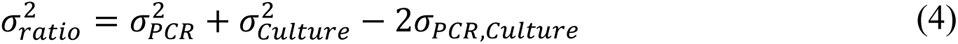

Here, *σ*_*PCR*,*Culture*_ is the covariance between qPCR and cultivation results on the log scale. The empirical covariance was estimated using the logarithm of the concentrations measured by qPCR and culture methods. By considering the covariance between the lognormal variables, Eq. 4 accounts for the dependency between the qPCR and culture results and adjusts the variance of the ratio accordingly.

Models were implemented using the Markov chain Monte Carlo (MCMC) method in a Bayesian framework. A uniform prior ranging from −10^2^ to 10^2^ and an exponential prior with a rate of 1.0 were specified for the location parameter μ and the shape parameter σ, respectively, for each lognormal distribution (qPCR and culture).

CFU and GC were estimated from reported concentrations, processed sample volumes, limits of quantification (LOQ) and limits of detection (LOD). The impact of analytical recovery rates on estimated concentrations was ignored due to insufficient reporting.

Cumulative distribution functions (CDFs) were used to illustrate distributions of *L. pneumophila* concentrations and qPCR:culture ratios, indicating the expected frequency of observing a ratio below a particular level. The best-fit curve was computed from the median values of the posterior distribution of each parameter. The uncertainty of the fit was represented with a 95% uncertainty interval obtained from the 2.5% and 97.5% percentiles of the posterior distribution of the parameters. The analysis was conducted in R (version 4.3.0). The R code used for data analysis and visualization can be found in the GitHub repository [NOTE: URL will be provided in the final paper].

### 3.2 Meta-analysis models

Statistical meta-analysis models were used to compare mean qPCR:culture ratios across multiple studies and obtain an overall distribution of the mean ratios. The geometric mean and arithmetic mean ratios were chosen as summary estimates, as they provide complementary information for interpreting the data. The mean log_10_ ratio, equivalent to the geometric mean ratio on the arithmetic scale, was chosen because it represents the median when the ratio is a lognormal random variable. Given the high skewness of qPCR:culture ratio distributions, the arithmetic mean was also computed for the meta-analysis. The importance of the arithmetic mean lies in its sensitivity to high ratios, which contribute to its value. This summary descriptor complements the geometric mean, which can suppress the impact of these high ratios. Results were illustrated using the *metafor* package version 3.0-2 (Viechtbauer, 2010) in R (version 4.3.0).

#### 3.2.1 Geometric mean ratio

Here, the meta-analysis is conducted on the mean log ratio on the natural log scale *μ*_ratio,1_, *μ*_*ratio*,2_, … *μ*_*ratio*,*k*_ and known standard errors of these means 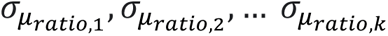. The location parameter of the lognormal distribution of the ratio is represented by *μ*_*ratio*_, and the standard error is given by:

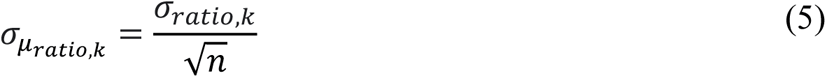

where *σ*_*ratio*,*k*_ is the scale parameter of the lognormal distribution of the ratio, and *n* is the sample size.

#### 3.2.2 Arithmetic mean ratio

In this case, the meta-analysis is conducted on the ratio *r̅*, which can be obtained from lognormal parameters as follows:

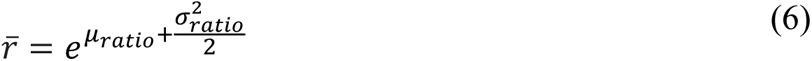

An estimator of the standard error (e.s.e.) of ln(*r̅*) can be derived following the approach presented by Olsson (2005). That is:

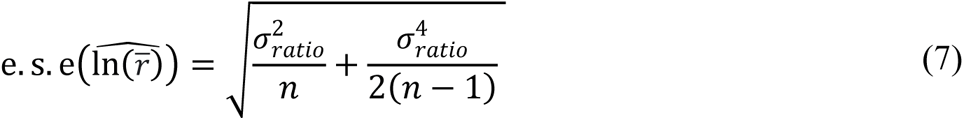

However, it is more convenient to express the e.s.e. of *r̅* on the log_10_-scale, as *r̅* is commonly presented in this format. Since *y* = −log_10_(*x*) is a continuous monotonic decreasing function, it follows that the e.s.e for log_10_(*r̅*) is:

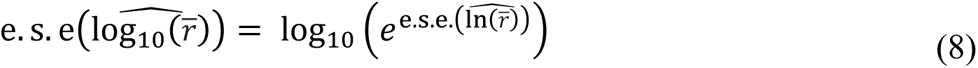

The proof of Eq. 8 is provided in Supplementary Material S2.

#### 3.2.3 Random-effects

A random-effects model assumes that the studies included in the meta-analysis are a random selection drawn from a broader pool of potential studies. These studies are assumed to be representative of a population describable by a single underlying distribution. This approach allowed us to account for both within-study and between-study variability. In the first stage of the model, the log_10_-transformed mean ratio (either geometric or arithmetic mean) from each study is given a weight based on its variance. These weights reflect the confidence we have in each set of observations of a study; the lower the variance, the higher the weight. Given the central limit theorem, the uncertainty around the estimated log mean ratio of each study is assumed to be normally distributed. For the geometric mean ratio of a study *k*, that is:

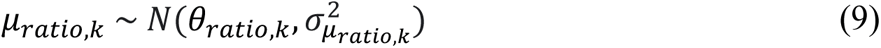

where *μ*_*ratio*,*k*_ is the inferred parameter of the parametric model of the ratio distribution (Eq. 3), *θ*_*ratio*,*k*_ is the true mean log ratio, and 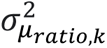 is the within-study variance (Eq. 5), representing the sampling uncertainty within each study.

The second stage of the model introduces a random effect to account for differences in log_10_-transformed mean ratios between studies. When we analyzed geometric mean ratios, we modelled this random effect with an exponential distribution. When we analyzed arithmetic mean ratios, we used a lognormal distribution. For a lognormal distribution with parameters *μ* and *τ*^2^, the second stage is:

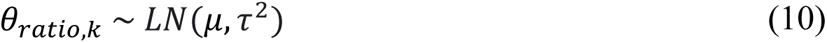

Here, *τ*^2^represents the between-study variance, capturing the variability in the true mean ratios (*θ*_*ratio*,*k*_) across studies. The parameters of these distributions were inferred with MCMC methods. We specified uninformative priors for each parameter, as proposed by Higgins et al. (2009). The exponential and the lognormal distributions were illustrated with CDFs.

#### 3.2.4 Subgroup analyses

Subgroup analyses were conducted to assess potential sources of variability in the mean ratios across various subgroups. The pooled estimates, representing the aggregated outcomes from each subgroup, were obtained using a Bayesian random-effects model. In this model, the within-study mean and the random effect were assumed to be normally distributed. Given the substantial uncertainty associated with subgroup pooled estimates due to small sample sizes, we focused on exploring the overall patterns and trends across the subgroups rather than conducting formal tests of significance between them.

## 4 Results

### 4.1 Data collection and reporting

Of the 1099 records screened, 189 full-text articles were assessed for eligibility, and 15 were selected for meta-analysis (Fig. 1). Ten of these studies reported the data from the studied locations as pooled, limiting our capacity to assess the influence of distinct water system characteristics and water quality parameters on *L. pneumophila* qPCR:culture ratios. Two studies (Donohue, 2021; Mapili et al., 2020) and five data sets from Lee et al. (2011) were excluded for having less than 20% quantifiable samples. All but two studies were conducted in European countries. One selected study, Lee et al. (2011), reported site-specific data from multiple potable water systems in different countries. Quality control and assurance elements reported for qPCR analyses varied among the 15 studies: one reported recovery rates for the whole process, four reported recovery rates for nucleic acid extraction, four reported doing positive/negative controls for nucleic acid extraction, 11 reported doing positive/negative controls for PCR detection, nine described methods for generating standard curves, and 12 reported using inhibition controls (Table S4). For cultivation analyses, all studies reported adhering to standardized protocols (Table S5), except two studies using Legiolert^®^ test, which were conducted before the publication of the ASTM D8429-21 standard in 2022.

**Fig. 1.**
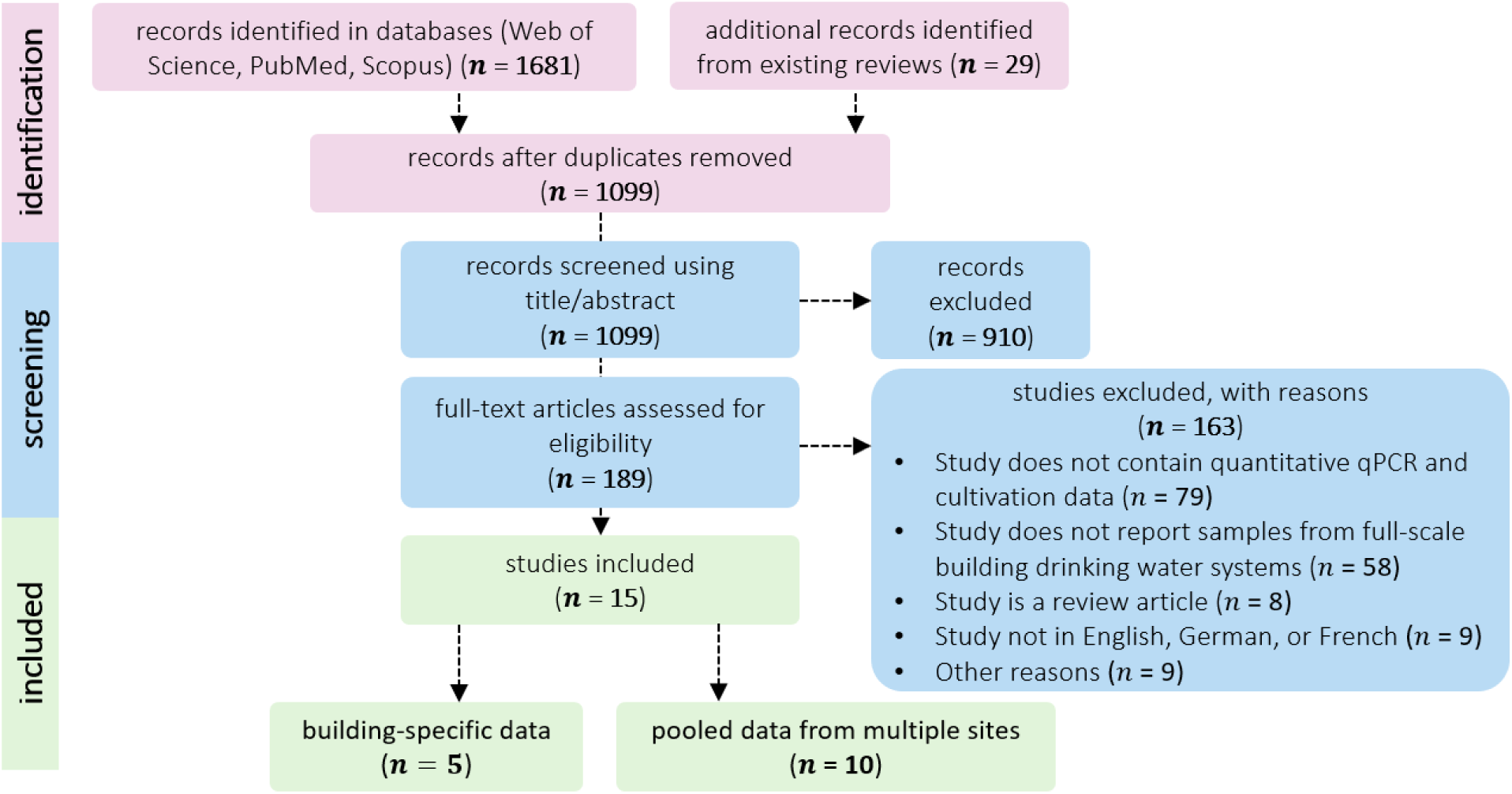
Systematic review flow diagram.

Several chemical disinfection strategies were used across studies, with the predominant approach being chlorine disinfection (Table 2). The water samples tested exhibited a range in average temperature of 22 to 57 °C. Sampling strategies vary significantly across studies. The number of sampling locations per building ranged from as few as one to as many as 21, with most studies monitoring less than five sampling locations. Approximately ten samples were typically collected at each sampling location, and most studies collected flushed samples (i.e., water has been intentionally run through the outlet being sampled before sampling). The volume analyzed for the quantification of *L. pneumophila* varied considerably, ranging from 27 mL to 1 L, for both culture-based and qPCR methods. The reported lower limit of detection ranged from 1 to 250 CFU L^-1^ or MPN L^-1^ for culture-based methods and 80 to 2000 GC L^-1^ for qPCR methods. Typically, the proportion of samples above the detection limit exceeded 70% when using qPCR. This proportion averaged 60% with culture-based methods, though this was as low as 20% for some sites.

**Table 2.**
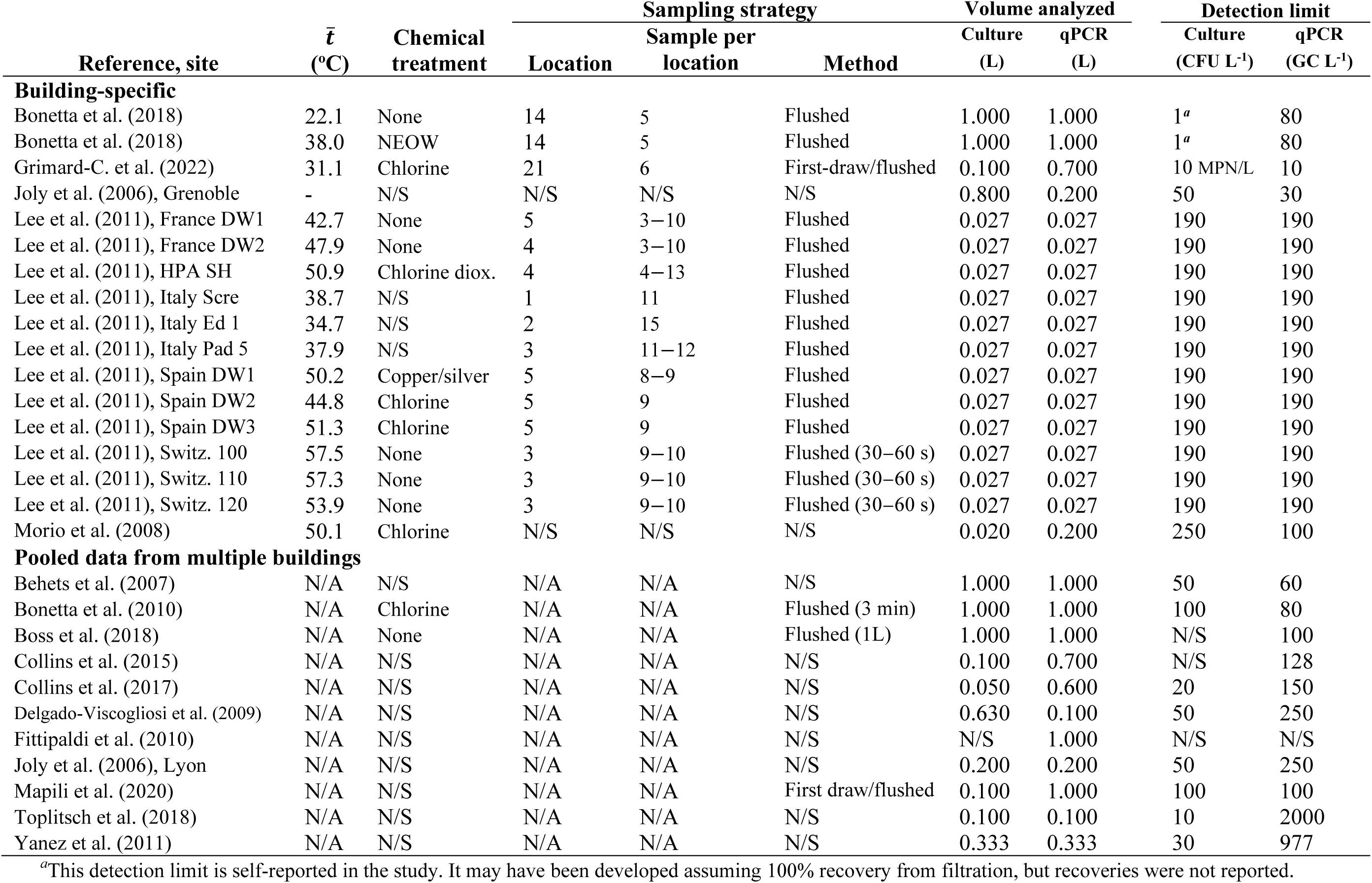
Characteristics of building potable water systems, sampling strategy and *Legionella pneumophila* enumeration methods for evaluating site-specific qPCR:culture ratio.

| Reference, site | $\bar{t}$<br>(°C) | Chemical<br>treatment | Sampling strategy | | | Volume analyzed | | Detection limit | |
| --- | --- | --- | --- | --- | --- | --- | --- | --- | --- |
|  |  |  | Location | Sample per<br>location | Method | Culture<br>(L) | qPCR<br>(L) | Culture<br>(CFU L<sup>-1</sup>) | qPCR<br>(GC L<sup>-1</sup>) |
| Building-specific |  |  |  |  |  |  |  |  |  |
| Bonetta et al. (2018) | 22.1 | None | 14 | 5 | Flushed | 1.000 | 1.000 | 1<sup>a</sup> | 80 |
| Bonetta et al. (2018) | 38.0 | NEOW | 14 | 5 | Flushed | 1.000 | 1.000 | 1<sup>a</sup> | 80 |
| Grimard-C. et al. (2022) | 31.1 | Chlorine | 21 | 6 | First-draw/flushed | 0.100 | 0.700 | 10 MPN/L | 10 |
| Joly et al. (2006), Grenoble | - | N/S | N/S | N/S | N/S | 0.800 | 0.200 | 50 | 30 |
| Lee et al. (2011), France DW1 | 42.7 | None | 5 | 3–10 | Flushed | 0.027 | 0.027 | 190 | 190 |
| Lee et al. (2011), France DW2 | 47.9 | None | 4 | 3–10 | Flushed | 0.027 | 0.027 | 190 | 190 |
| Lee et al. (2011), HPA SH | 50.9 | Chlorine diox. | 4 | 4–13 | Flushed | 0.027 | 0.027 | 190 | 190 |
| Lee et al. (2011), Italy Scre | 38.7 | N/S | 1 | 11 | Flushed | 0.027 | 0.027 | 190 | 190 |
| Lee et al. (2011), Italy Ed 1 | 34.7 | N/S | 2 | 15 | Flushed | 0.027 | 0.027 | 190 | 190 |
| Lee et al. (2011), Italy Pad 5 | 37.9 | N/S | 3 | 11–12 | Flushed | 0.027 | 0.027 | 190 | 190 |
| Lee et al. (2011), Spain DW1 | 50.2 | Copper/silver | 5 | 8–9 | Flushed | 0.027 | 0.027 | 190 | 190 |
| Lee et al. (2011), Spain DW2 | 44.8 | Chlorine | 5 | 9 | Flushed | 0.027 | 0.027 | 190 | 190 |
| Lee et al. (2011), Spain DW3 | 51.3 | Chlorine | 5 | 9 | Flushed | 0.027 | 0.027 | 190 | 190 |
| Lee et al. (2011), Switz. 100 | 57.5 | None | 3 | 9–10 | Flushed (30–60 s) | 0.027 | 0.027 | 190 | 190 |
| Lee et al. (2011), Switz. 110 | 57.3 | None | 3 | 9–10 | Flushed (30–60 s) | 0.027 | 0.027 | 190 | 190 |
| Lee et al. (2011), Switz. 120 | 53.9 | None | 3 | 9–10 | Flushed (30–60 s) | 0.027 | 0.027 | 190 | 190 |
| Morio et al. (2008) | 50.1 | Chlorine | N/S | N/S | N/S | 0.020 | 0.200 | 250 | 100 |
| Pooled data from multiple buildings |  |  |  |  |  |  |  |  |  |
| Behets et al. (2007) | N/A | N/S | N/A | N/A | N/S | 1.000 | 1.000 | 50 | 60 |
| Bonetta et al. (2010) | N/A | Chlorine | N/A | N/A | Flushed (3 min) | 1.000 | 1.000 | 100 | 80 |
| Boss et al. (2018) | N/A | None | N/A | N/A | Flushed (1L) | 1.000 | 1.000 | N/S | 100 |
| Collins et al. (2015) | N/A | N/S | N/A | N/A | N/S | 0.100 | 0.700 | N/S | 128 |
| Collins et al. (2017) | N/A | N/S | N/A | N/A | N/S | 0.050 | 0.600 | 20 | 150 |
| Delgado-Viscogliosi et al. (2009) | N/A | N/S | N/A | N/A | N/S | 0.630 | 0.100 | 50 | 250 |
| Fittipaldi et al. (2010) | N/A | N/S | N/A | N/A | N/S | N/S | 1.000 | N/S | N/S |
| Joly et al. (2006), Lyon | N/A | N/S | N/A | N/A | N/S | 0.200 | 0.200 | 50 | 250 |
| Mapili et al. (2020) | N/A | N/S | N/A | N/A | First draw/flushed | 0.100 | 1.000 | 100 | 100 |
| Toplitsch et al. (2018) | N/A | N/S | N/A | N/A | N/S | 0.100 | 0.100 | 10 | 2000 |
| Yanez et al. (2011) | N/A | N/S | N/A | N/A | N/S | 0.333 | 0.333 | 30 | 977 |

<sup>a</sup>This detection limit is self-reported in the study. It may have been developed assuming 100% recovery from filtration, but recoveries were not reported.

### 4.2 Temporal variations in qPCR:culture ratios

We observed substantial variability in the qPCR:culture ratio, with most sites demonstrating ratios spanning an approximate 3.0-log (Figure 2). For every data set, the lower tail of the lognormal distribution of the ratio began to rise within a qPCR:culture ratio of 0.1 to 1.0. Theoretically, ratios below 1.0 are unexpected since the number of genome copies detected by qPCR should be equal to or higher than the number of culturable cells. However, the model predicts such low ratios when data sets include multiple cultivation and qPCR results that fall below the detection limits. The upper tail of the distribution varied among studies. Most sites exhibited maximum ratios between 1.0 and 2.0-log, while a few presented maximum ratios exceeding 3.0-log. In many studies, a small number of samples with high qPCR:culture ratios considerably influenced the arithmetic mean ratio (e.g., Bonetta et al. (2018), Grimard-Conea et al. (2022), Lee et al. (2011); Table 3).

**Fig. 2.**
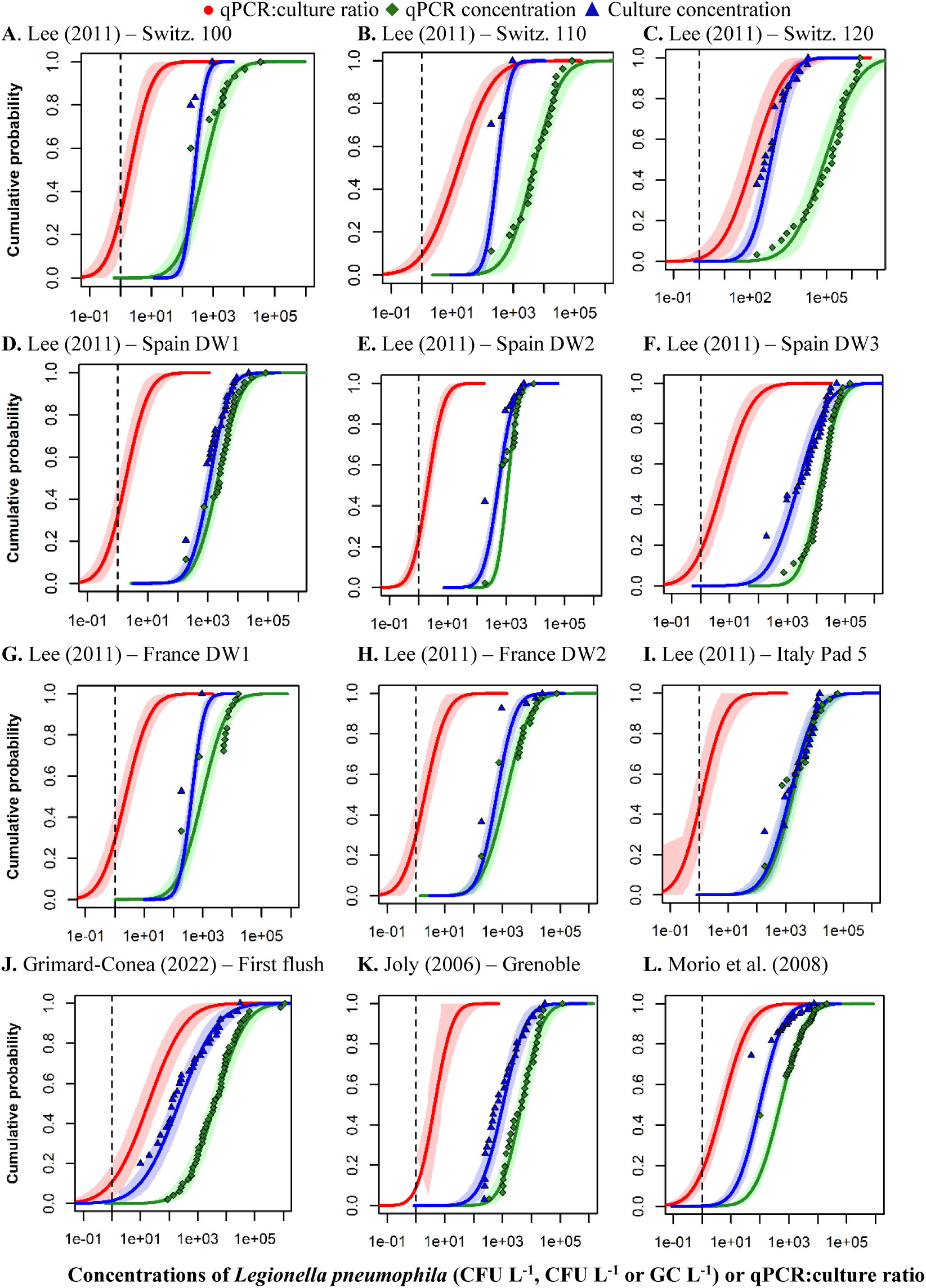
Complementary cumulative distribution function of the lognormal distribution (with 95% uncertainty interval) for building-specific qPCR and culture concentrations of *Legionella pneumophila* and their ratio in water samples from selected building potable water systems. Only 12 of the 17 sites are shown to illustrate main trends.

**Table 3.**
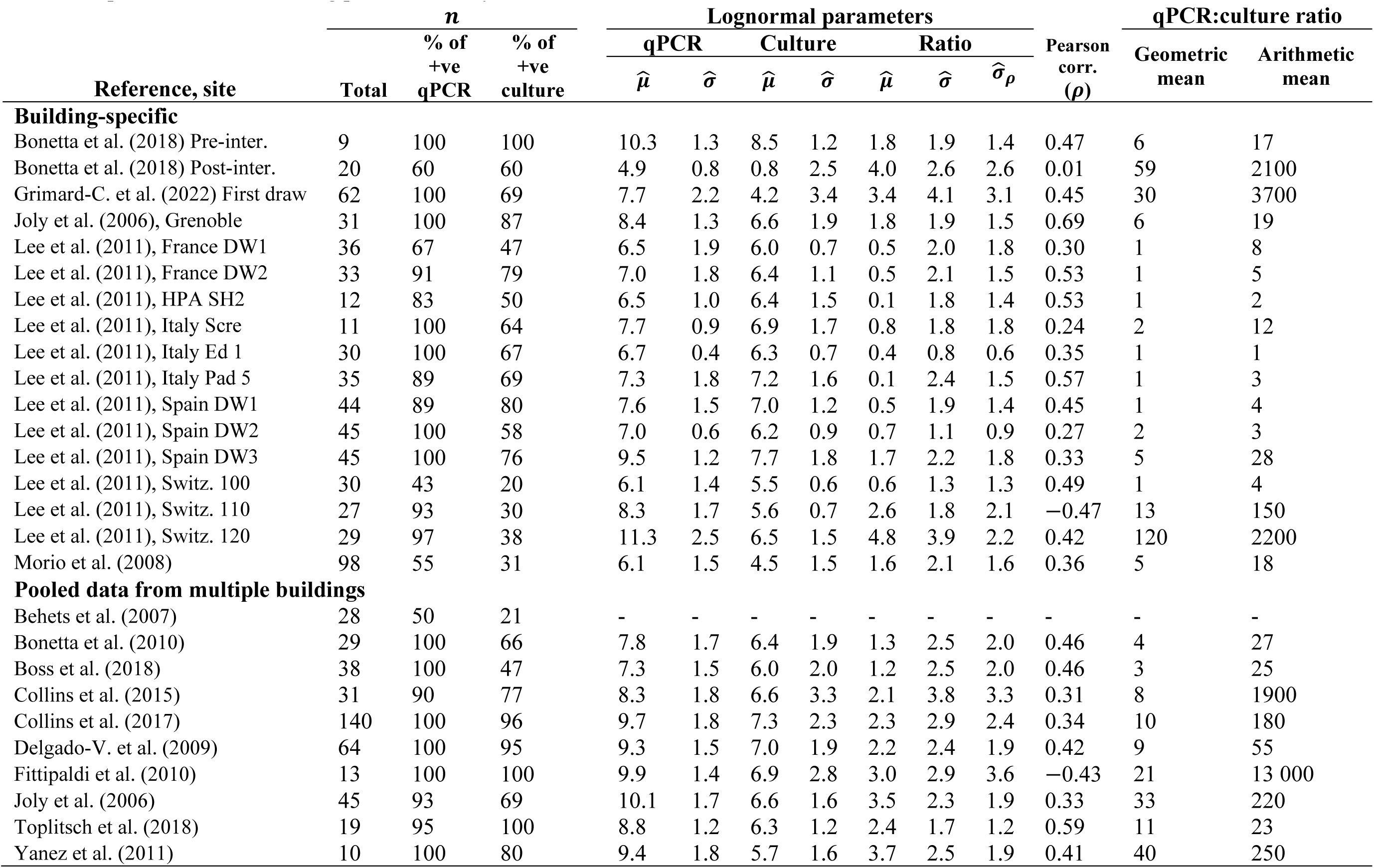
Statistical analysis of site-specific qPCR:culture ratios, qPCR concentrations and culture concentrations for *Legionella pneumophila* in water samples collected in building potable water systems.

This analysis demonstrates that the distribution of the concentration—either derived from qPCR or culture-based measurements—can dominate the dispersion of the ratio distribution (Figure 2). When culture-based measurements returned a high number of non-detects, the qPCR concentration distribution predominantly influenced the distribution of the ratio (Fig 2A, 2B, 2G). In other cases, the culture-based concentration distribution dictated the distribution of the ratio (Fig 2F, 2J). As shown in Table 3, the correlation coefficient also affects the dispersion of the ratio distribution. This finding highlights the need to account for the interdependence between the two methods to characterize the ratio distribution accurately.

### 4.3 Between-study variability in mean qPCR:culture ratios

For reviewed studies reporting site-specific data, the geometric mean qPCR:culture ratios display considerable variability, ranging from 0.0 to 2.1-log (Fig. 3). Exponential and lognormal distributions of the random effect illustrate how the geometric or arithmetic mean ratios varies across studies (Fig. 4). The exponential distribution predicts geometric mean qPCR:culture ratios below 10:1 (1.0-log) for approximately 80% of the sites. This suggested that qPCR and cultivation results were similar in most locations. For the remaining 20% of the sites, the geometric mean ratio was predicted to fall between 1.0 and 2.0-log. The distribution of arithmetic mean ratios displayed more skewed, with 50% of the sites presenting mean ratios above 1.0-log and considerable uncertainty surrounding the distribution, indicating that high ratios were expected at these sites.

**Fig. 3.**
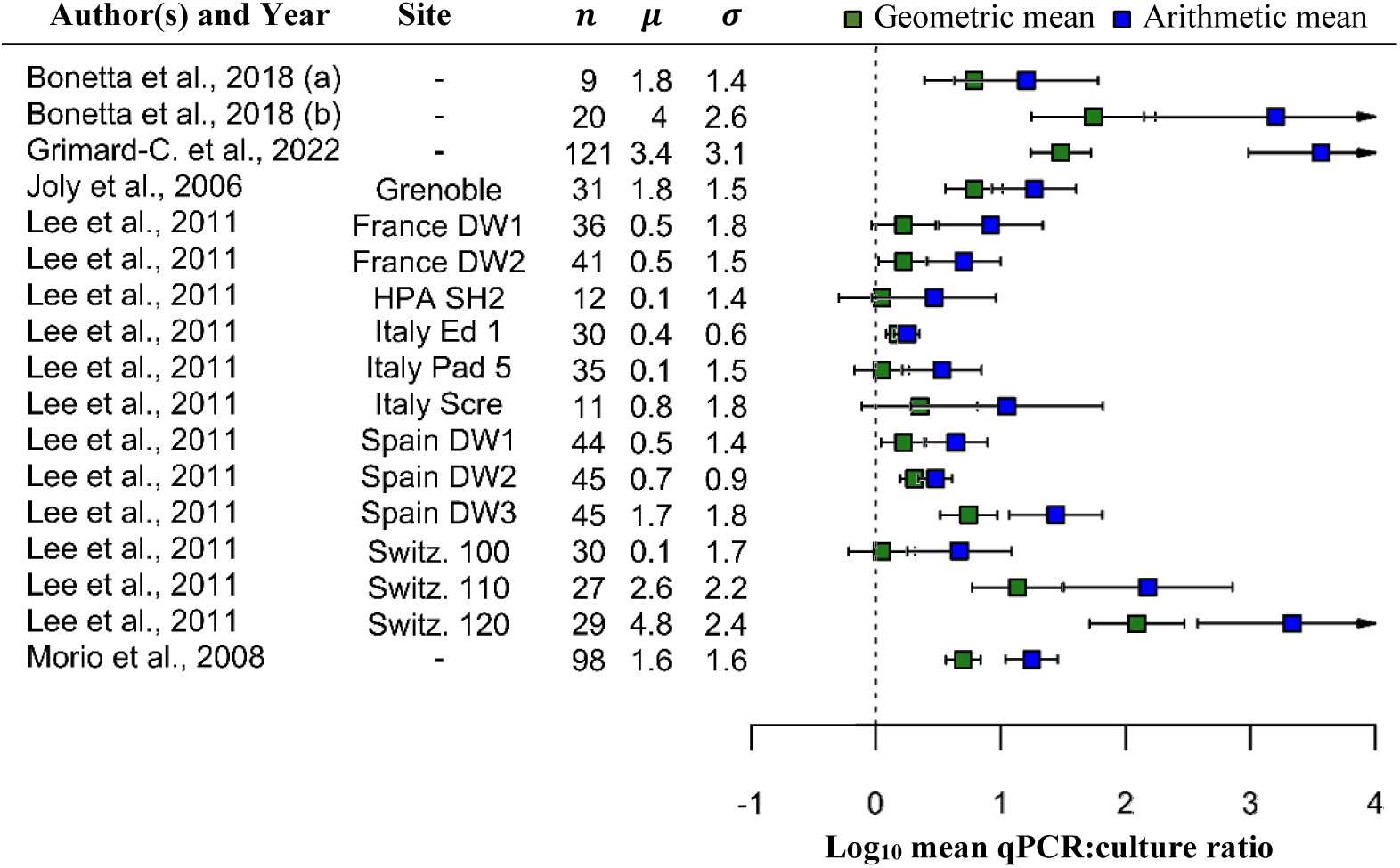
Forest plots of site-specific qPCR:culture ratios for *Legionella pneumophila* in building potable water systems for arithmetic mean ratios and geometric mean ratios. *μ* and *σ* are the parameters of the lognormal distribution of the ratio, and *n* is the sample size. Horizontal lines represent 95% confidence intervals on mean values. Arrows indicate that confidence intervals exceed a log-ratio value of 4.0.

**Fig. 4.**
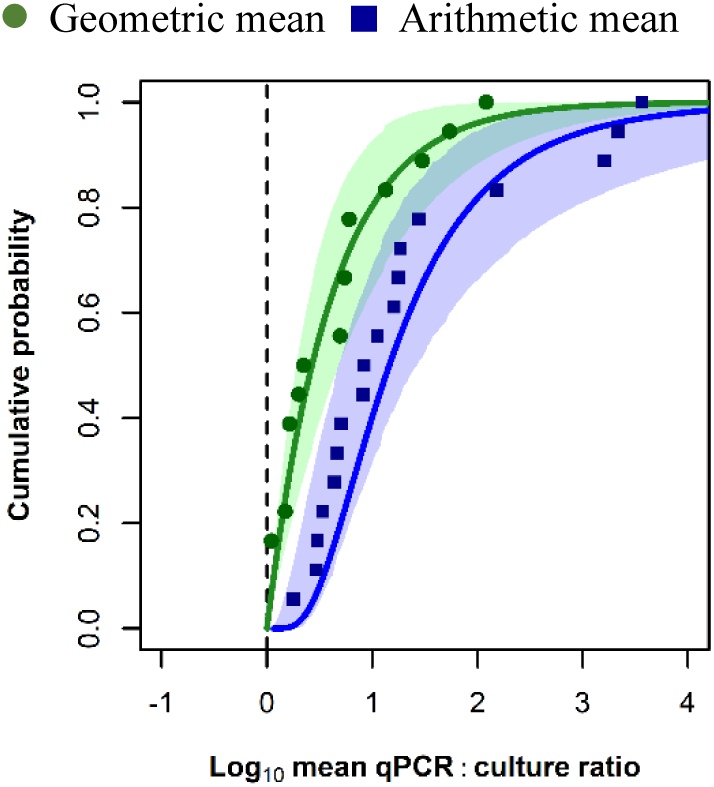
Cumulative distribution function of the random-effects distribution (with 95% uncertainty interval) predicted using an exponential random-effects model for geometric mean qPCR:culture ratios (green) and a lognormal random-effects model for arithmetic mean qPCR:culture ratios (blue) for *Legionella pneumophila* in building potable water systems.

### 4.4 Impact of viability-qPCR on qPCR:culture ratios

For the three studies employing viability-qPCR, arithmetic and geometric mean qPCR:culture ratios display lower values than standard qPCR (Fig. 5, Fig. S2, Table S3). Across all studies, viability-qPCR reduced geometric mean ratios by 0.2-log and arithmetic mean ratios by 0.5-log. For the studies conducted by Bonetta et al. (2018) and Yáñez et al. (2011), arithmetic mean ratios were more uncertain for standard qPCR than viability-qPCR, suggesting that viability-qPCR can reduce the variability in ratios.

**Fig. 5.**
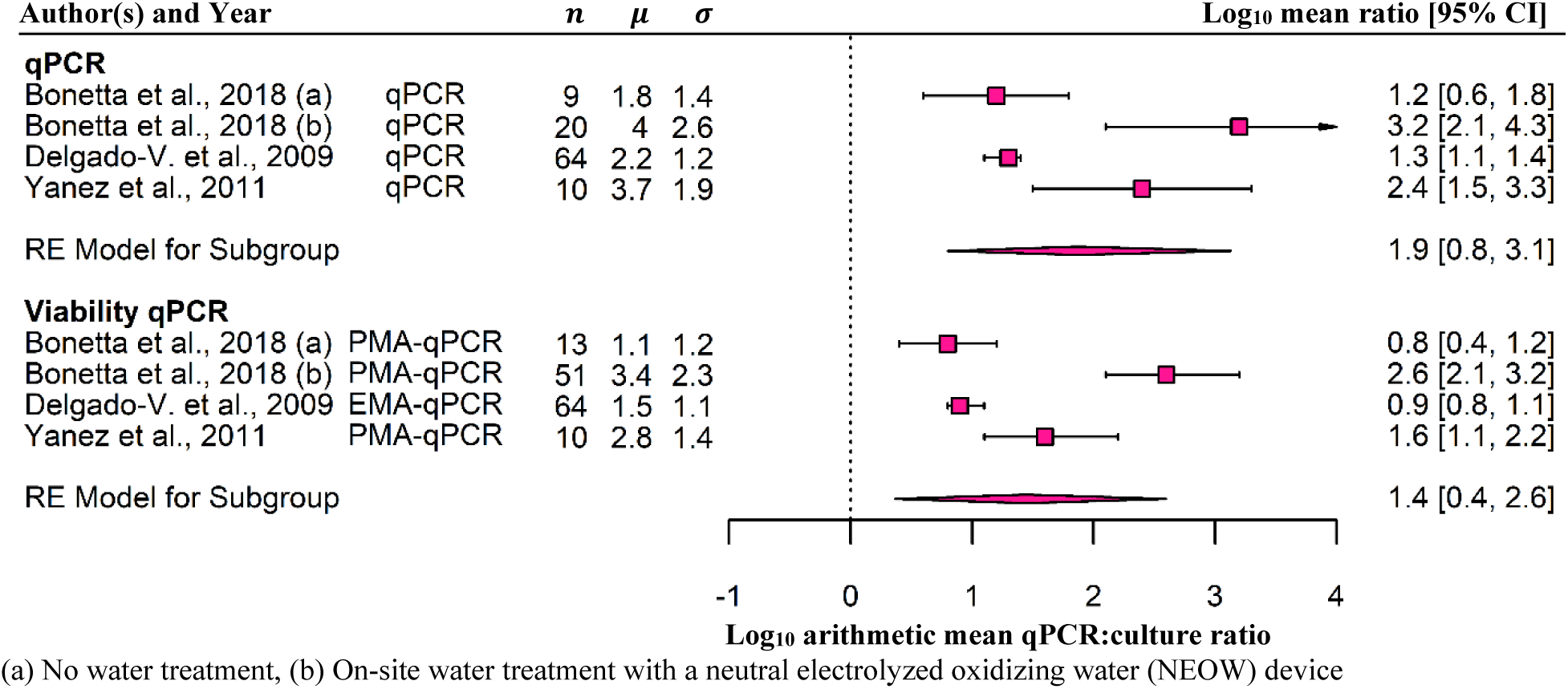
Forest plots of qPCR:culture ratio and viability-qPCR:culture ratio for *Legionella pneumophila* in water samples from building potable water systems. Subgroup analyses were carried out for arithmetic mean ratios. *μ* and *σ* are the parameters of the lognormal distribution of the ratio, and *n* is the sample size. Horizontal lines represent 95% confidence intervals on mean values. Pooled estimates were obtained using the random-effect model.

### 4.5 Impact of water temperature on qPCR:culture ratios

The analysis of impacts of temperature on qPCR:culture data was limited to one study (Lee et al.), as this was the only study that reported sample-specific temperature data for each study site. The influence of hot water temperature on qPCR:culture ratios was examined using data sets from sites Switzerland 100, 110 and 120. During the monitoring period, no chemical residuals, on-site chemical treatment, or shock disinfection were applied at these three sites (V. Gaia, personal communication, April 2023). qPCR:culture ratios increased appreciably across the three sites as the temperature became more variable with intermittently lower temperatures, as shown in the CDFs (Fig. 6). At site 100, the water temperature from 30−60 sec flush samples consistently stayed above 55 °C. Conversely, the other sites recorded lower temperatures, with values as low as 47 °C at site 110 and 32 °C at site 120.

**Fig. 6.**
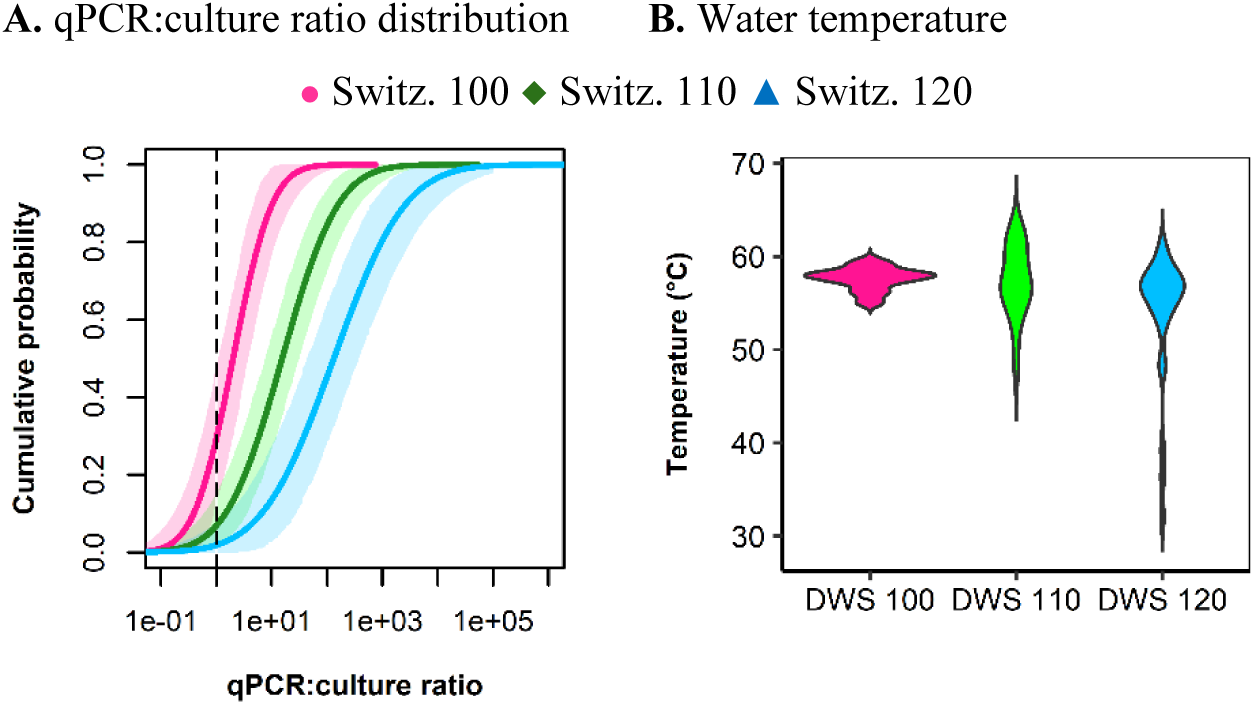
**A.** Cumulative distribution function of the lognormal distribution (with 95% uncertainty interval) for qPCR:culture ratios of *Legionella pneumophila* in flushed water samples from non-chlorinated building potable water systems. Samples were collected weekly for ten weeks at three different sampling locations for each water system. **B.** Violin plots of the water temperature based on sample-specific temperature measurements.

## 5 Discussion

This review considered the complex interplay between qPCR and culture-based methods by modelling published data sets reporting *L. pneumophila* concentrations measured in building potable water systems. We found that, predominantly, qPCR yields higher measurements than culture-based methods, a difference influenced by several factors. Higher measurements with qPCR compared to cultivation likely arise from the detection of uncultivable cells and extracellular DNA (Delgado-Viscogliosi et al., 2009). The detection of all genetic material could overestimate health risks, placing an unnecessary burden on building managers. Conversely, conventional culture-based methods may underestimate risks due to cultivation bias, or by not adequately quantifying bacterial aggregates or VBNC cells, even though thermally induced VBNC *L. pneumophila* can have lower virulence compared to culturable *L. pneumophila* (Cervero-Aragó et al., 2019). This divergence between qPCR and cultivation results may further be influenced by analytical strategies and environmental variables (Lee et al., 2011), including water temperature, disinfectant levels, and water stagnation, which in turn can be affected by operational parameters like boiler settings as well as flushing and sampling strategies. Given this complexity, caution is necessary when converting qPCR data for use in QMRA.

### 5.1 Ecology of *Legionella pneumophila* in building potable water systems

The temporal analysis of qPCR:culture ratios within building potable water systems shows that while some water samples have comparable concentrations in both qPCR and cultivation, others exhibit much higher concentrations when measured by qPCR compared to cultivation. Typically, the temporal distribution is right-skewed with a long tail towards high ratios, suggesting that a small proportion of samples is characterized by high concentrations of *L. pneumophila* nucleic acids not associated with culturable bacteria. The water temperature is one potential explanation for variability in qPCR:culture ratios, as high water temperatures may reduce culturability while minimally influencing qPCR detection. Sample-specific temperature data from Lee et al. (2011) enabled us to investigate how temperature impacts variations in qPCR:culture ratios in non-chlorinated hot water systems from three hospitals in Switzerland. At site Switzerland 110, qPCR concentrations displayed high variations of approximately 3.0-log, while culture concentrations were predominantly at the LOD. These results suggest that *L. pneumophila* enter the hot-water heater through recirculation lines or potentially grow within the water heater but are then inactivated by the thermal barrier in the water heater, hence a high qPCR:culture ratio. In contrast, site Switzerland 120 demonstrated high variability in qPCR concentrations, but multiple culture-positive results were recorded. This observation points toward potential *L. pneumophila* growth in the hot-water distribution system or a compromised disinfection capacity of the hot-water heater. This hypothesis is supported by water sample temperatures measured at this site, which sporadically fell within the ideal temperature for *L. pneumophila* growth (25−43°C) (National Academies of Sciences and Medicine, 2020). Notably, a full-scale study by Bédard et al. (2016) demonstrated that implementing corrective measures to maintain a water temperature of 55 °C throughout the hot-water system of a large building resulted in a gradual reduction in concentrations of *L. pneumophila*, as measured by both qPCR and cultivation.

Curative control measures, such as thermal and chemical shock treatments, can impact qPCR:culture ratios. In the full-scale study conducted by Grimard-Conea and Prévost (2023), published after our review period, shock chlorination combined with flushing regimes resulted in culturable *L. pneumophila* concentrations below the LOD, while qPCR still indicated concentrations of approximately 10^3^ genome-copies per liter. Additionally, under bench-scale conditions, Delgado-Viscogliosi et al. (2009) demonstrated that exposing *L. pneumophila* to 70°C water for 60 min reduced cultivability by more than 6.0-log. In contrast, nucleic acid integrity was only affected by approximately 0.2-log. Despite clear evidence that such measures impact ratios, there were an insufficient number of studies to provide conclusive quantitative estimates of the impact of remediation strategies on qPCR:culture ratios.

### 5.2 qPCR data integration into *L. pneumophila* risk assessment

A conversion from genome-copies to CFU is necessary for QMRA because available *L. pneumophila* dose–response models predict health risks for inhaled doses in CFU (Armstrong and Haas, 2007; Armstrong and Haas, 2008). To integrate qPCR data into QMRA, previous studies converted genome copies to an estimate of viable pathogen counts using a constant qPCR:culture ratio (McBride et al., 2013). This approach can be effective if qPCR:culture ratios remain relatively stable over time. However, a stable ratio may not be likely for *L. pneumophila* in building potable water systems. These systems provide variable conditions, with possible inactivation of *L. pneumophila* in a point-of-entry treatment system (e.g., hot-water heater) but also possible (re)growth in the plumbing system before the point of use.

Our meta-analysis indicates that qPCR:culture ratios tend to be low and exhibit significant variability. For about 80% of the building potable water systems, predicted geometric mean qPCR:culture ratios were less than 10, with predicted ratios approaching 1:1 at all sites. Therefore, to avoid underestimating the concentration of viable *L. pneumophila*, applying a 1:1 adjustment factor appears necessary when converting a *L. pneumophila* qPCR result into cultivation result to quantify *L. pneumophila* concentrations in building potable water for QMRA. Using higher constant qPCR:culture ratios like 10:1, 100:1, or 1000:1 to adjust monitoring results could mask *L. pneumophila* growth events, thereby underestimating health risks associated with these systems. This recommendation applies exclusively to *L. pneumophila*; this review did not target other *Legionella* species. While some conservative risk assessments might apply *L. pneumophila* dose–response models for other *Legionella* species, the interpretation of health risks associated with *Legionella* species is currently limited due to the absence of dose–response models for species beyond *L. pneumophila* (Armstrong and Haas, 2007; Armstrong and Haas, 2008).

The recommended 1:1 factor might significantly overestimate health risks in scenarios where point-of-entry treatment systems, such as thermal disinfection or on-site chemical disinfection, reduce the culturability or viability of *L. pneumophilia*, with minimal-to-no impact on DNA concentrations. Directly using qPCR concentrations to estimate risks in such systems may result in an excessive number of sites being erroneously classified as hazardous, triggering unnecessary interventions and imposing undue burdens on facility managers. However, determining an adjustment factor that accurately captures the impact of disinfection from point-of-entry treatment systems is challenging. This difficulty arises because *L. pneumophila* can growth within a hot-water line post-disinfection or at a fixture, potentially leading to lower qPCR:culture ratios compared to those measured directly from the effluent of the hot-water heater. Consequently, relying on a qPCR:culture ratio based solely on the inactivation efficacy of the point-of-entry treatment system to adjust qPCR data for QMRA may mask *L. pneumophila* growth events. More accurate estimates of potential health risks in such systems could be obtained by pairing measurements from qPCR methods with those from culture-based or viability-qPCR methods. Therefore, qPCR should, at this stage, be viewed as a complementary tool for risk assessment and building investigations, rather than as a replacement for cultivation.

Data on *L. pneumophila* concentrations derived from qPCR can provide useful insights into water safety above and beyond what is possible using culture-data alone. For example, elevated or sudden increases in qPCR-based concentrations can infer *L. pneumophila* growth within the system. This information can become valuable when considered within the context of the site. For instance, it can highlight areas requiring intervention such as disinfection, elimination of dead ends, improvement in temperature controls, maintenance, or redesign. For population segments more vulnerable to developing severe Legionnaires’ disease, including the elderly, immunocompromised individuals, smokers, and those with chronic lung conditions, qPCR results may be valuable for identifying high-risk systems.

Viability-qPCR can reduce and stabilize qPCR:culture ratios by excluding non-viable cells (Lizana et al., 2017; Yáñez et al., 2011). The subgroup analysis conducted in this study revealed that, on average, viability-qPCR leads to a reduction of approximately 0.2-log in the geometric mean and 0.5-log in the arithmetic mean when compared to standard qPCR. This reduction appears relatively low, considering the range of mean ratios expected from the meta-analysis. A partial explanation is that the viability-qPCR methods are conservative (i.e. only targeting extensively damaged cells) (Hammes et al., 2011). Nonetheless, targeted uses of viability-qPCR in specific scenarios, such as the effluent of a contaminated hot-water heater with a sufficient thermal barrier, could provide valuable insights. By employing viability-qPCR in these circumstances, it could become possible to determine whether the overwhelming majority of *L. pneumophila* detected by standard qPCR are non-viable. Further research into the conditions under which VBNC *L. pneumophila* can regain virulence and the factors influencing their transition back to a culturable state could also be beneficial to improve risk assessment accuracy.

### 5.3 Improved reporting in qPCR:culture comparison studies

The lack of comprehensive data reporting limited our ability to accurately quantify uncertainties associated with concentrations of *L. pneumophila* measured by qPCR and culture-based methods. In all studies, only estimated concentrations were reported. However, for modelling temporal variations in pathogen concentrations, it is preferable to use original lab observations (counts and processed volumes). This approach allows for incorporating statistical uncertainties into the analysis (Chik et al., 2018; Schmidt et al., 2023). In our study, we estimated cultivation counts from reported concentrations and sample volumes, but this method can underestimate uncertainties when samples undergo dilution or subsampling. To accurately back-calculate counts from reported cultivation concentrations, it is necessary to have access to dilution factors and volume plated for plating methods, positive and negative well counts for MPN methods, and elution and template volumes for qPCR methods. LODs from qPCR and culture-based methods in reviewed studies sometimes differed by orders of magnitude. Minimizing differences between LODs would be beneficial as it would enhance the accuracy of qPCR:culture ratios at low concentrations. Ideally, when qPCR results are positive, corresponding culturable concentrations should be detectable. More pairing of positive results could increase the covariance between qPCR and cultivation, reducing the variability in the ratio distribution (see Eq. 4). This adjustment might correct for the tendency of the lower tail of the ratio distribution to predict ratios below 1.0, which may facilitate the adoption of less conservative conversion factor than 1:1 in some situations.

Comparing qPCR and cultivation data can be further complicated by differences in analytical recovery rates between the two methods. However, we were unable to evaluate this impact due to insufficient reporting. Whole process recovery rates for the cultivation of *L. pneumophila* in drinking water samples have been found to range from 30–90% for direct plating (Blanky et al., 2015; Boulanger and Edelstein, 1995; Villari et al., 1998) and 12–48% for pre-treated samples (Blanky et al., 2015; Boulanger and Edelstein, 1995). For qPCR, reported recovery rates have ranged from 42–98% for the whole process (Behets et al., 2007; Omiccioli et al., 2015) and 57– 123% specifically for DNA extraction (Collins et al., 2015; Collins et al., 2017). Although the direct impact of these variations on the qPCR:culture ratio may be relatively minor compared to certain environmental factors, the effect of this methodological factor remains uncertain and warrants further investigation.

On a broader scale, while the study by Lee et al. (2011) adhered to the AFNOR NF-T90-471:2010 standard for qPCR testing, other studies reviewed did not follow an established qPCR standard. Many studies only partially reported specific controls and reporting elements recommended in the Minimum Information for Publication of Quantitative Real-Time PCR Experiments (MIQE) guidelines (Bustin et al., 2009) or the Environmental Microbiology Minimum Information (EMMI) guidelines (Borchardt et al., 2021). Adopting these guidelines in future research would improve the reliability and reproducibility of qPCR results.

Without standardized methods for both cultivation and qPCR, consistent correlations between qPCR and cultivation concentrations in building potable water systems remain uncertain (Whiley and Taylor, 2014). However, even with standardized methods, variations in environmental factors such as water temperature can still impact culturability and affect correlations in site-specific temporal analyses. To quantify the influence of these environmental variables on the relationship, mathematical models, such as qPCR:culture ratio distributions and multiple regression models can be applied, provided that site-specific contextual data are reported. The parameters listed in Table 1 offer a preliminary framework for more standardized reporting.

## 6 Conclusions

This meta-analysis examined the qPCR-to-culture concentration ratio for *L. pneumophila* in building potable water systems, aiming to guide the interpretation of qPCR data for QMRA. The key conclusions drawn from this study are as follows:

- Our analysis confirms that Poisson lognormal distributions effectively describe system-specific variations in concentrations of *L. pneumophila* using both qPCR and culture-based methods.
- The relationship between qPCR and culture methods, modeled as the ratio of two correlated lognormal random variables, showed substantial variability within systems, with ratios typically ranging from 1:1 to 100:1. This variability can be attributed to environmental factors such as water temperature and methodological considerations, including detection limits.
- Across the literature, the random-effects meta-analysis model indicated that for approximately 80% of systems, the geometric mean of the qPCR-to-culture ratios is less than 10:1. However, due to occasional high ratios, the arithmetic mean ratios exceed 100:1 in about 20% of the systems.
- Given the observed variability and frequent occurrences of ratios close to 1:1, implementing a default 1:1 conversion factor for converting qPCR data to culturable concentrations in QMRA is recommended. This strategy avoids underestimating culturable concentrations due to regrowth of *L. pneumophila* downstream of point-of-entry treatment systems like hot water heaters.
- In cases of heavy *L. pneumophila* contamination before point-of-entry treatment systems, directly incorporating qPCR data into QMRA might overestimate health risks by detecting large amounts of DNA from non-infectious bacteria. Emerging techniques like viability-qPCR, which targets DNA within intact cells, could provide a more accurate alignment of concentration estimates with actual health risks associated with *L. pneumophila*. However, these techniques are not yet widely adopted in qPCR-based applications.
- More standardized, site-specific reporting can facilitate detailed analysis of environmental and methodological variables influencing qPCR-to-culture ratios. This includes studying conditions that are likely to yield high proportions of non-viable and viable but non-culturable (VBNC) *L. pneumophila*.

## Data Availability

All data produced are available online at: https://tinyurl.com/44tx6a5s

## 7 Acknowledgments

ES was funded by postdoctoral fellowships from the Natural Sciences and Engineering Research Council of Canada (558161-2021) and the Fonds de Recherche du Québec Nature et technologies (303866). Additional funding was provided by the Swiss Federal Food Safety and Veterinary Office (FSVO), the Federal Office of Public Health (FOPH) and the Federal Office of Energy (FOE), through the project LeCo (Legionella Control in Buildings; Aramis nr.: 4.20.01), as well as Eawag discretionary funding. We gratefully acknowledge Nadine Graf for her assistance with data extraction and compilation, Brittany Halverson-Duncan and Prof. Peter Vanrolleghem for their guidance on statistical analyses, and Dr. Valeria Gaia and Dr. John V. Lee for providing data and sharing their insights for interpreting the findings.

## List of symbols

38 List of symbols
|  |  |
| --- | --- |
| $c$ | *Legionella pneumophila* concentration |
| $k$ | Individual study |
| $n$ | Sample size |
| $Q$ | Chi-square statistic |
| $\bar{r}$ | Arithmetic mean qPCR:culture ratio on the raw scale |
| $V$ | Sample volume |
| $x$ | *Legionella pneumophila* count |
| $\mu$ | Mean parameter of the lognormal distribution |
| $\mu_{Culture}$ | Mean parameter of the lognormal distribution of culture concentrations |
| $\mu_{PCR}$ | Mean parameter of the lognormal distribution of qPCR concentrations |
| $\mu_{ratio}$ | Mean parameter of the lognormal distribution of qPCR:culture ratios |
| $\theta_{ratio}$ | True mean log qPCR:culture ratio |
| $\sigma$ | Standard deviation parameter of the lognormal distribution |
| $\sigma_{Culture}$ | Standard deviation parameter of the lognormal distribution of culture concentrations |
| $\sigma_{PCR}$ | Standard deviation parameter of the lognormal distribution of qPCR concentrations |
| $\sigma_{Ratio}$ | Standard deviation parameter of the lognormal distribution of qPCR:culture ratios |
| $\sigma_{PCR,Culture}$ | Covariance between qPCR and culture results on the log scale |
| $\sigma_{\mu_{ratio}}$ | Standard error of $\mu_{ratio}$ |
| $\sigma_{\theta_{ratio}}$ | Standard error of $\theta_{ratio}$ |
| $\tau^2$ | Between-study variance of $\theta_{ratio}$ |
| $e.s.e(\widehat{\ln(\bar{r})})$ | Estimator of the standard error (e.s.e.) of $\ln(\bar{r})$ |
| $e.s.e(\widehat{\log_{10}(\bar{r})})$ | Estimator of the standard error (e.s.e.) of $\bar{r}$ on the $\log_{10}$ scale |

## Notes

### Competing Interest Statement

The authors have declared no competing interest.

