## Supplemental Material for "Quantification of *Legionella pneumophila* in building potable water systems: a meta-analysis comparing qPCR and culture-based detection methods"

**Supplementary Information Contents:**

**S1 Protocol for the systematic review and meta-analysis**

Protocol outlining the detailed steps and methods for conducting our systematic review and meta-analysis, including search strategy, selection criteria, data extraction, and statistical methods.

**S2 Mathematical proof**

Proof for the transformation of the estimated standard error of the arithmetic mean on the log_10_ scale.

**S3 Additional results**

Additional figures and tables not included in the main paper.

**S1 Protocol for the systematic review and meta-analysis**

This protocol is based on the 2015 PRISMA-P guideline (1, 2).

### 1 Administrative information

#### 1.1 Title

**Identification:** Method comparability for *Legionella* measurements in building potable water systems: a protocol for systematic review and meta-analysis.

**Update**: Version 1, May 2022

#### 1.2 Registration

This protocol was not registered.

#### 1.3 Contributors

***Contact***

Corresponding author: Émile Sylvestre

Author Affiliation

Department of environmental microbiology, Swiss Federal Institute of Aquatic Science and Technology (Eawag), Überlandstrasse 133 8600 Dübendorf, Switzerland.

***Contributions***

ES is the guarantor. ES and WR drafted the manuscript and contributed to developing the selection criteria, the risk of bias assessment strategy and the data extraction criteria. WR developed the search strategy. ES provided statistical expertise. All authors provided expertise on *Legionella*. All authors read, provided feedback and approved the final manuscript.

#### 1.4 Amendments

Amendments of the protocol will be described and documented in Appendix A. The report will be provided as supplementary material for the final article.

#### 1.5 Support

***Sources***

This research is funded by the Federal Food Safety and Veterinary Office (FSVO), in partnership with the Federal Offices of Public Health (FOPH) and Energy (SFOE) in Switzerland, through the project LeCo (Legionella Control in Buildings; Aramis nr.:4.20.01) and Eawag discretionary funding. ES is supported by postdoctoral fellowships from the Natural Sciences and Engineering Research Council of Canada (NSERC) and the Fonds de Recherche du Québec - Nature et Technologies (FRQNT).

***Sponsor***

N/A

### 2 Introduction

#### 2.1 Rationale

Inhalation of aerosols containing the bacterium *Legionella pneumophila* can cause illness in humans (legionellosis), including a pneumonia-type illness known as Legionnaires’ disease and a mild flu-like illness known as Pontiac fever. Legionella is commonly found in aerosols produced showers, faucets, spa pools, fountains, cooling towers and other engineered water systems. To prevent legionellosis, the concentration of *Legionella* in the bulk water of a system generating these aerosols needs to be controlled at an acceptable level, which can be determined using quantitative microbial risk assessment (QMRA) (Figure 1) (3). Validated monitoring strategies and accurate enumeration methods are therefore required to be able to determine whether an acceptable concentration is exceeded or not.

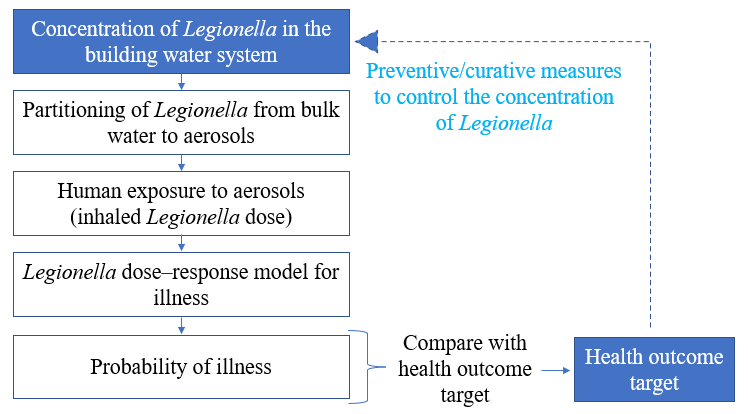

**Fig. 1**. QMRA approach for defining risk-based control measures to meet a health outcome target (adapted from WHO (4))

Monitoring of water samples for *Legionella* is coming increasingly into use as a result of the development and standardization of culture-based methods (ISO 11731:2017; NF T90-431:2017; ASTM D8429-21) and quantitative polymerase chain reaction (qPCR)-based methods (ISO/TS 12869:2019). With qPCR, quantitative results (number of genome copies [GC] per unit volume) can be obtained in a few hours, which is much faster than the several days required to obtain quantitative results with culture (number of colonies forming units [CFU] per unit volume). However, in contrast with culture, qPCR does not quantify viable *Legionella* cells only; this molecular method captures all DNA: viable, dead and viable but non-culturable (VBNC) cells. Other factors can explain differences in results obtained with the two methods, including variable analytical recovery rates for both techniques, method-specific limits of detection, and potential bias in culture due to aggregation of *Legionella* cells in water samples. Interpretation of qPCR-generated results in terms of human health risk is not straightfoward because available dose–reponse models for *L. pneumophila* predict risks for average doses measured with culture (CFU) (5). Harmonizing qPCR and culture measurements is needed to incorporate qPCR data in risk assessment models.

Several method comparison studies investigating the relationship between concentrations of *Legionella* measured in water samples by qPCR and culture methods have been published since 2000. These studies indicate that detection rates and concentrations are typically higher with qPCR than with culture; however, differences in concentrations obtained with both methods can vary by orders of magnitude within and between water systems (6). Further investigation of the heterogeneities in the qPCR/culture concentration relationship is warranted to help interpret qPCR results, particularly how preventive and curative control strategies influence the relationship. This work is critical to ensuring risk managers can make informed decisions regarding the selection and use of methods for the enumeration of *Legionella* in water samples.

#### 2.2 Objectives

The aim of this systematic review and the meta-analysis is to compare *Legionella* measurements evaluated with qPCR and culture-based methods to facilitate the interpretation of qPCR-generated data. To this end, the following research question was formulated considering two elements (Population, Outcomes) of the PECO framework (7).

“What is the ratio between concentrations of *Legionella* (*L. pneumophila* or *L.* species) measured with culture methods (ISO 11731, Legiolert) and PCR methods (qPCR, viability qPCR) (O) in water samples collected in full-scale building potable water systems (P)?”

### 3 Methods

#### 3.1 Eligibility criteria

Studies will be selected according to the criteria outlined below:

- The Population is water samples collected in full-scale building potable water systems. Bench-scale and pilot-scale studies will be excluded from the review.
- The Outcome (quantity of interest) is the ratio between concentrations of *Legionella* (*L. pneumophila* or *L.* species) measured with culture methods (ISO 11731, Legiolert) and PCR methods (qPCR, viability qPCR) in individual samples. Other PCR methods, such as digital droplet PCR (ddPCR) and reverse-transcription PCR (RT-qPCR), will be excluded from the review.
- A preliminary search showed that PCR assays for the quantitative determination of *Legionella* in water samples were published from 2000 onwards; therefore, studies published before 2000 will be excluded.
- The study was published in a peer-reviewed journal or is a governmental report. Review, commentaries, letters, editorials, conference proceedings, and posters will be excluded.
- The search will be limited to English, French and German.

#### 3.2 Information sources

Backward and forward searches have already been conducted by one reviewer, starting with a narrative review paper on methods comparison for *Legionella* enumeration (8) and the National Academies of Science, Engineering and Medicine (NASEM) report Management of Legionella in Water Systems (3). This strategy revealed that about 30 method comparison studies had been published since 2000. Results from this search strategy will be used to inform the electronic search strategy. Studies published in peer-reviewed journals will be searched in PubMed, Scopus, and Web of Science. Governmental reports in English or German will be identified via the network of researchers and stakeholders of the project LeCo. A data request email template will be used to contact study authors when relevant data cannot be extracted from studies.

#### 3.3 Search strategy

The search string for PubMed, Scopus, and Web of Science is the following:

“(Legio* AND qPCR AND culture) OR (Legio* AND ddPCR AND culture) OR (Legio* AND viability PCR AND culture) OR (Legio* AND EMA AND culture) OR (Legio* AND PMA AND culture) OR (Legio* AND Methods AND Compare)”

#### 3.4 Study records

***Data management***

Google sheets will be used to retrieve citations, screen citations, and record reasons for exclusion. All data extracted from studies will also be stored in Google Sheets to facilitate collaborative document editing.

***Selection process***

Titles and abstracts will be screened for potentially relevant studies using a “liberal accelerated” approach, i.e., potentially relevant citations are identified by one reviewer, and a second reviewer verifies potential excluded citations. Two independent reviewers will complete the full-text screening, and any discrepancies will be resolved through discussion. This stage will be carried out based on the following questions:

1. Does the abstract refer to primary research reported in a peered-review journal publication (as opposed to a review article)?
2. Were samples of water collected from full-scale building potable water systems?
3. Were concentrations of *L. pneumophila* or *L.* species quantified by qPCR and culture-based methods?

Studies will be included in the review only if the answers to each question are “Yes”. Reasons for excluding studies will be recorded.

***Data collection process***

Single data extraction will be carried out. Water system information, sample processing information, measurement information, data reporting information and concentrations (see section 3.5) will be extracted by two reviewers and verified by a third reviewer. Reported concentrations will be extracted from tables. WebPlotDigitizer (<https://automeris.io/WebPlotDigitizer/>) will be used to extract approximated concentrations from figures when concentrations are not reported in tables. Study authors will be contacted to resolve any uncertainties. Data requests for raw databases will be sent to study authors (maximum of three email attempts) when concentrations data cannot be extracted from tables or figures.

#### 3.5 Data items

Data items extracted in each publication are listed in Table 1.

**Table 1.** Parameter to be extracted from each study

| **Category** | **Parameter to extract** |
| --- | --- |
| Water system information | - Building type - Water system - Outlet - Draw - Geographical location |
| Water treatment | - Preventive control measure(s) - Curative control measure(s) |
| Water quality parameter | - Total chlorine - Free chlorine - Monochloramine - Water temperature |
| Sample processing information | - Sample type - Sample volume for each method - Volume processed for each method |
| Measurement information | - Limits of detection for each method - Limit of quantification for each method |
| Data reporting information | - Data pooling strategy |
| *Legionella* quantification | - Concentration of *L.* spp. for each method - Concentration of *L. pneumophila* for each method |

#### 3.6 Outcomes and prioritization

N/A

#### 3.7 Risk of bias in individual studies

The risk of bias (or internal validity) describes the extent to which limitations in the design and conduct of the study can affect the interpretation of concentrations of *L. pneumophila* or *L*. spp. Bias domain and assessment questions for evaluating the internal validity of individual studies are listed in Table 2, following an adaptation from the Cochrane Collaboration Risk of Bias Tool (9) and D. Rodríguez-Molina et al. (10). Risks of bias will be described as low risk/high risk/unclear risk to inform data synthesis. Two reviewers will evaluate the risk of bias in selected studies, and any discrepancies will be resolved through discussion.

**Table 2**. Risk of bias assessment

| **Bias domain** | **Assessment question** | **Criteria** |
| --- | --- | --- |
| Measurement bias | “Were enumeration methods used in a way that ensures the same internal validity regardless of the collected sample?” | 1. Criteria for the judgement of “Yes”:   - Identical methods applied to all samples - Controlling for laboratory factors, if these are different   2. Criteria for the judgement of “No”:   - Application of different methods depending on the water system; - No adjustment strategy for different laboratory methods   3. Risk of bias will be considered “unclear” if there is not enough information to judge information bias criteria as either “yes” or “no”, e.g. if enumeration methods are not explained sufficiently to reach a judgement |
| Information bias | “Were site-specific water systems information available to interpret results from samples collected?” | 1. Criteria for the judgement of “Yes”:   - Sample-specific concentrations are reported for each water system   2. Criteria for the judgement of “No”:   - Only pooled data from multiple systems are reported   3. Risk of bias will be considered “unclear” if there is not enough information to judge information bias criteria as either “yes” or “no”, e.g. if the water system information is not explained sufficiently to reach a judgement |
| Information bias | “Were curative control measures potentially affecting concentrations reported adequately?” | 1. Criteria for the judgement of “Yes”:   - The date and time of curative control measures are reported (if any)   2. Criteria for the judgement of “No”:   - Curative control measures are reported but without date and time   3. Risk of bias will be considered “unclear” if there is not enough information to judge information bias criteria as either “yes” or “no”, e.g. if control measures strategies are not explained sufficiently to reach a judgement |

#### 3.8 Data synthesis

***Method for summarising data from an individual study***

The qPCR/culture concentration ratio will be calculated from the extracted data. All sample-specific ratios with a non-detect in culture or qPCR will be excluded from the analysis. The impact of this choice will be evaluated with a sensitivity analysis. The ratio data will be transformed on the log_10_-scale to reduce the skewness of its distribution. The arithmetic mean ratio on the logarithmic scale, or equivalently, the geometric mean ratio on the raw scale will be evaluated from individual studies. The arithmetic mean ratio on the raw scale would be a preferable descriptor to summarize site-specific data for QMRA (11). However, preliminary work shows that most studies report pooled data, not site-specific data. In this situation, the geometric mean on the raw scale is a preferable summary measure to describe the central tendency of the pooled data.

The ratio data will be assumed to be lognormally distributed. It has been demonstrated that this approach is reasonably robust to different skewed distributions (12). To estimate the mean log-ratio, let $X$ denote a random variable with a lognormal distribution, then $Y=log(X)$ has a normal distribution with parameters $\mu$ and $\sigma$. The mean $\mu$ and variance $\sigma^{2}$ of the normal distribution can be estimated with minimum variance unbiased estimators as follows:

$$\hat{\mu}=\bar{x}=\frac{1}{n}\sum_{i=1}^{n} x_{i}$$

(1)

$$\hat{\sigma}^{2}=s^{2}=\frac{1}{n-1}\sum_{i=1}^{n} \left( x_{i}-\bar{x} \right)^{2}$$

(2)

(13). These estimates will be computed from raw data using the function *elnorm* from the R package EnvStats (14).

The random error, quantifying the statistical precision of the ratio, will be evaluated by calculating the $\left( 1-\alpha\right)100\%$ confidence interval for the mean log-ratio $\hat{\mu}$ as follows:

| $[\hat{\mu}-t\left( n-1, 1-\alpha/2 \right)\frac{\hat{\sigma}}{\sqrt{n}}, \hat{\mu}+t\left( n-1, 1-\alpha/2 \right)\frac{\hat{\sigma}}{\sqrt{n}}]$ | (3) |
| --- | --- |

where $t(\upsilon,\rho)$ is the $\rho$’th quantile of Student’s t-distribution with $\upsilon$ degrees of freedom (15). The $\left( 1-\alpha\right)100\%$ confidence interval for the variance $\hat{\sigma}^{2}$ is given by:

| $[\frac{\left( n-1 \right)s^{2}}{\chi_{n-1, 1-\alpha/2}^{2}}, \frac{\left( n-1 \right)s^{2}}{\chi_{n-1, \alpha/2}^{2}}]$ | (4) |
| --- | --- |

(16). A minimum sample size requirement will be established to ensure that the estimated variance $\hat{\sigma}^{2}$ is reasonably accurate.

***Criteria under which study data will be quantitatively synthesized***

The combined use of qualitative and quantitative methods will be used to synthesize results from multiple studies. During the early stage of the data synthesis, results from large groups of studies will be synthesized. These initial groups may be refined as the data synthesis develops. Any amount of heterogeneity will be acceptable if predefined eligibility criteria are met. Substantial heterogeneity is expected in a meta-analysis of the ratio of concentrations of microorganisms in environmental water, and multiple meta-analysis methods account for between-studies heterogeneities (17). The challenge will be to find appropriate covariates and data sets to investigate possible causes of the between-study heterogeneity.

***Quantitative synthesis***

Inverse-variance weighting will be used to weight mean ratios from individual studies. This method minimizes the variance of the weighted average by weighting each mean proportionally to its precision (i.e., the inverse of its variance) (18). Therefore, if within-study means $\hat{\mu}_{1},\ldots, \hat{\mu}_{k}$ from $k$ studies are independent and have respective within-study estimated variances $\hat{\sigma}_{1},\ldots, \hat{\sigma}_{2}$, then the variance of the weighted mean is minimized by the optimal weight:

$$\hat{w}_{i}=\frac{1}{\hat{\sigma}_{i}^{2}}$$

(5)

If optimal weights are normalized such that $\sum_{i}^{k} \hat{w}_{i}=1$, then the weight is given by

$$\hat{w}_{i}^{*}=\frac{1}{\hat{\sigma}_{i}^{2}}/\sum_{i=1}^{k} \frac{1}{\hat{\sigma}_{i}^{2}}$$

(6)

A fixed-effect model will be used to estimate the common mean from $k$ studies. Suppose that a set of $\hat{\mu}_{1},\ldots, \hat{\mu}_{k}$ within-study means and that optimal weights $\hat{w}_{1}^{*},\ldots, \hat{w}_{k}^{*}$ are normalized using Eq. 6. The common mean estimated with the fixed-effect model is given by

$$\hat{\mu}_{Pooled}=\sum_{i=1}^{k} \hat{w}_{i}^{*}\hat{\mu}_{i}$$

(7)

The variance of $\hat{\mu}_{FE}$ will be estimated using the method M. Henmi and J. B. Copas (19). This method was selected because substantial between-study heterogeneity is expected. The variance is given by:

$$Var(\hat{\mu}_{Pooled})=\sum_{i=1}^{k} \left[ \left( \hat{w}_{i}^{*} \right)^{2}(\hat{\sigma}_{i}^{2}+\hat{\tau}^{2}) \right]$$

(8)

where $\tau^{2}$ is the estimated between-study variance.

The meta-analysis results will be visually explored with forest plots. This $I^{2}$ index will be used to measure heterogeneity in the meta-analysis. This index describes the percentage of variation across studies due to heterogeneity rather than sampling error. Heterogeneity will be categorized as low, moderate and high at 25%, 50% and 75%, respectively.

The meta-analysis will be conducted in the statistical software environment R using the *metafor* package version 3.0-2 (20).

***Qualitative synthesis***

Given the diversity in building types, water systems, and water treatment, it is likely that some data sets will not be appropriate for statistical meta-analyses. A systematic qualitative (narrative) synthesis will be undertaken in such cases. This narrative synthesis will include textual descriptions of studies, a table that summarizes the results, and thematic analyses, in line with the Guidance on the conduct and reporting of narrative synthesis from the Economic and Social Research Council (ESRC) (21). Tables will be used to summarize (a) the mean log-ratio of studies and (b) the characteristics of studies. Priority will be given to water system characteristics and water treatment characteristics. Studies with a high risk of bias will be included in the qualitative synthesis only if they provide the available data for thematic analyses.

***Subgroup analyses***

Subgroup analyses will be used to explore possible sources of heterogeneity based on the following.

1. Water system information;
2. Water quality parameters;
3. Water treatment.

The method of M. Henmi and J. B. Copas (19) will be used to calculate pooled estimates from each group. Differences between subgroups will be evaluated by observing how the confidence interval of the pooled estimate overlap.

***Sensitivity analyses***

Sensitivity analysis will be performed to explore the source of heterogeneity as follows:

- The impact of the risk of bias will be evaluated by conducting the same meta-analysis but omitting studies that are judged to be at high risk of bias.
- The impact of excluding all sample-specific ratios with a non-detect in culture or qPCR will be evaluated. Sample-specific ratios with a non-detect in culture will be calculated assuming that the non-detect has a concentration equal to the limit of detection reported in the studies. The same statistical models will be used to calculate individual study estimates and pooled estimates.

3.	National Academies of Sciences E, Medicine. 2020. Management of Legionella in water systems. National Academies Press.

4.	WHO. 2016. Quantitative microbial risk assessment: Application for water safety management. Geneva, Switzerland.

13.	Johnson NL, Kotz S, Balakrishnan N. 1995. Continuous univariate distributions, volume 2, vol 289. John wiley & sons.

14.	Millard SP, Kowarik A, Kowarik MA. 2018. Package ‘EnvStats’. Packag Environ Stat Version 2:31-32.

15.	Helsel DR, Hirsch RM. 1992. Statistical methods in water resources, vol 49. Elsevier.

16.	Zar JH. 1999. Biostatistical analysis. Pearson Education India.

24.	Olsson U. 2005. Confidence intervals for the mean of a log-normal distribution. Journal of Statistics Education 13.

25.	Higgins JP, Thompson SG, Spiegelhalter DJ. 2009. A re‐evaluation of random‐effects meta‐analysis. Journal of the Royal Statistical Society: Series A (Statistics in Society) 172:137-159.

**Appendix A: Amendments to the original protocol**

---------------------------------------------------------------------------------------------------------------------

**Amendment #1: Modification of the risk of bias assessment questions**

The original protocol proposes a methodology for analyzing the risk of bias in the design and conduct of studies. The assessment questions were modified as follows:

1. To evaluate the risk of measurement bias, the question

“Were enumeration methods used in a way that ensures the same internal validity regardless of the collected sample?”

was changed to

“Is the paper referencing the Minimum Information for Publication of Quantitative Real-Time PCR Experiments (MIQE) guidelines or the Environmental Microbiology Minimum Information (EMMI) guidelines?

This information was reported in the Discussion Section of the main paper.

2. To evaluate the risk of information bias, the question

“Were site-specific water systems information available to interpret results from samples collected?”

was changed to

“Were site-specific *Legionella pneumophila* data reported?”

Results were used to build Table 1 of the main paper.

3. To evaluate the risk of information bias, the question

“Were curative control measures potentially affecting concentrations reported adequately?”

was changed to

“Were water temperatures and chemical treatments reported?”

Results were included in Table 1 of the main paper.

---------------------------------------------------------------------------------------------------------------------

**Amendment #2: Refinement of the original statistical methods for quantitative synthesis**

Our initial protocol proposed to use standard meta-analysis methods to summarize data from individual stud and to perform a quantitative synthesis of literature data. We significantly enhanced these methods in the course of our research.

To better analyze the qPCR:culture ratios derived from individual studies, we used mixed Poisson models. This approach allowed us to handle non-detects and model the variability more accurately. We also adapted our meta-analysis models to assess the between-study variability in both the geometric and arithmetic mean qPCR:culture ratios. This change provides an improved understanding of the differences in ratio distributions across studies.

Our updated statistical approach is outlined below.

#### Variability of the qPCR:culture ratio within a building

To estimate how much the ratio between the results of the two methods (qPCR and culture) varies within each building, we used a statistical method previously described by É. Sylvestre et al. (22). This method involves comparing two sets of results that follow a specific probability distribution, known as a Poisson$-$lognormal distribution. To apply this model, the counts $x$ (CFU for culture and GC for qPCR) are assumed to be randomly distributed in each water sample of volume $V$ and concentration $c$. The Poisson distribution states that the probability of finding $x$ organisms is:

$$P\left( x;c,V \right)=\frac{\left( cV \right)^{x}e^{-cV}}{x!}$$

(1)

Organisms have a concentration $c$, thus their expected number in a sample volume $V$ is $cV$. The concentration $c$ is expected to vary in time or space. This variation can be described by a lognormal distribution. The probability density function of the lognormal distribution is:

$$f\left( c;\mu,\sigma\right)=\frac{1}{c\sigma\sqrt{2\pi}}\exp\left( -\frac{\left( \ln\left( c \right)-\mu\right)^{2}}{2\sigma^{2}} \right)$$

(2)

where $\mu$ and $\sigma$ are, respectively, the mean and the standard deviation of the underlying normal distribution on the logarithmic scale. If Eq. 2 describes the variation of $c$ in Eq. 1, then the count, $x$, follows a Poisson$-$lognormal distribution. The quality of the fit of the Poisson$-$lognormal (PLN) distribution was compared to the quality of the fit of an alternative model, the Poisson$-$gamma (PGA) distribution, using the marginal deviance information criterion (mDIC) (23). Preliminary results indicated that the lognormal distribution better described concentration variations for qPCR data (Table S1) and culture data (Table S2). Consequently, the Poisson$-$lognormal distribution was chosen for modelling *L. pneumophila* concentrations and their ratios.

The distribution of the ratio of two lognormal random variables is also lognormally distributed with a mean of:

$$\mu_{ratio}=\mu_{PCR}-\mu_{Culture}$$

(3)

and a variance of:

$$\sigma_{ratio}^{2}=\sigma_{PCR}^{2}+\sigma_{Culture}^{2}-2\sigma_{PCR, Culture}$$

(4)

Here, $\sigma_{PCR, Culture}$ represents the covariance between PCR and culture results on the log scale. The empirical covariance was estimated using the logarithm of the concentrations measured by qPCR and culture methods. By considering the covariance between the lognormal variables, Eq. 4 accounts for the dependency between the qPCR and culture results and adjusts the variance of the ratio accordingly.

The PLN and PGA distributions were implemented using the Markov chain Monte Carlo (MCMC) method in a Bayesian framework. A uniform prior ranging from $-$10^2^ to 10^2^ and an exponential prior with a rate of 1.0 were specified for the location parameter μ and the shape parameter σ, respectively, for each lognormal distribution (qPCR and culture).

CFU and GU were estimated from reported concentrations, processed sample volumes, limits of quantification (LOQ) and limits of detection (LOD). The count was set to zero for results by culture-based methods at the LOD. For results by qPCR, counts at the LOD and LOQ were computed assuming concentrations were equal to the LOQ and LOD. Only data sets with at least 20% of quantifiable samples were modelled. Analytical recovery rates likely vary by method. For this analysis, the impact was ignored due to insufficient reporting.

Cumulative distribution functions (CDFs) were used to illustrate distributions of *L. pneumophila* concentrations and qPCR:culture ratios, indicating the expected frequency of observing a ratio below a particular level. The best-fit curve was computed from the median values of the posterior distribution of each parameter. The uncertainty of the fit was represented with a 95% uncertainty interval obtained from the 2.5% and 97.5% percentiles of the posterior distribution of the parameters. The analysis was conducted in R (version 4.3.0). The R code for the analysis is available in the Supplementary Material.

#### Meta-analysis models

Meta-analysis models were used to compare mean qPCR:culture ratios across multiple studies and obtain an overall distribution of the mean ratios. For the meta-analysis, both the geometric mean ratio and arithmetic mean ratio were chosen as summary estimates, as they provide complementary information for interpreting the data. The mean log_10_ ratio, equivalent to the geometric mean ratio on the arithmetic scale, was chosen because it represents the middle ground value of the results (i.e., the median) when the ratio is a lognormal random variable. Given the high skewness of qPCR:culture ratio distributions, the arithmetic mean was also computed for the meta-analysis. The importance of the arithmetic mean lies in its sensitivity to high ratios, which contribute to its value. This summary descriptor complements the geometric mean, which can suppress the impact of these high ratios.

##### *Geometric mean ratio*

Here, the meta-analysis is conducted on the mean log ratio on the natural log scale $\mu_{ratio,1},\mu_{ratio,2},\ldots\mu_{ratio,k}$ and known standard errors of these means $\sigma_{\mu_{ratio,1}},\sigma_{\mu_{ratio,2}},\ldots\sigma_{\mu_{ratio,k}}$. The location parameter of the lognormal distribution of the ratio is represented by $\mu_{ratio}$, and the standard error is given by:

$$\sigma_{\mu_{ratio,k}}=\frac{\sigma_{ratio,k}}{\sqrt{n}}$$

(5)

where $\sigma_{ratio,k}$ is the scale parameter of the lognormal distribution of the ratio, and $n$ is the sample size.

##### *Arithmetic mean ratio*

In this case, the meta-analysis is conducted on the ratio $\bar{r}$, which is obtained by applying a formula that converts the lognormal mean and variance to the arithmetic mean on the exponential scale as follows:

$$\bar{r}=e^{\mu_{ratio}+\frac{\sigma_{ratio}^{2}}{2}}$$

(6)

An estimator of the standard error (e.s.e.) of $\ln(\bar{r})$ can be derived following the approach presented by U. Olsson (24). That is:

$$e.s.e\left( \hat{\ln\left( \bar{r} \right)} \right)=\sqrt{\frac{\sigma_{ratio}^{2}}{n}+\frac{\sigma_{ratio}^{4}}{2\left( n-1 \right)}}$$

(7)

However, it is more convenient to express the e.s.e. of $\bar{r}$ on the log_10_-scale, as $\bar{r}$ is commonly presented in this format. Since $y=-\log_{10} \left( x \right)$ is a continuous monotonic decreasing function, it follows that the e.s.e for $\log_{10} \left( \bar{r} \right)$ is:

$$e.s.e\left( \hat{\log_{10} \left( \bar{r} \right)} \right)= \log_{10} \left( e^{e.s.e.\left( \hat{\ln\left( \bar{r} \right)} \right)} \right)$$

(8)

The proof of Eq. 8 is provided in the Supplementary Material.

##### Random-effects

A random-effects model is a specific type of model that assumes that the studies included in the analysis are a random sample from a larger population. This meta-analysis model considers both within and between-study variability.

In the first stage of the model, the mean ratio (either geometric or arithmetic mean) from each study is given a weight based on its variance. These weights reflect the confidence we have in each set of observations of a study; the lower the variance, the higher the weight. We assumed that the observed mean ratio of each study follows a normal distribution. For the geometric mean ratio of a study $k$, that is:

$$\mu_{ratio, k} \sim N(\theta_{ratio, k}, \sigma_{\mu_{ratio,k}}^{2})$$

(9)

where $\mu_{ratio, k}$ is the inferred parameter of the parametric model of the ratio distribution (Eq. 3), $\theta_{ratio, k}$ is the true mean log ratio, and $\sigma_{\mu_{ratio,k}}^{2}$ is the within-study variance (Eq. 5), representing the sampling uncertainty within each study.

The second stage of the model introduces a random effect to account for differences in mean ratios between studies. Depending on whether we are analyzing geometric or arithmetic mean ratios, we modelled this random effect with either an exponential or lognormal distribution. For a lognormal distribution with parameters $\mu$ and $\tau^{2}$, the second stage is:

$$\theta_{ratio, k} \sim LN(\mu, \tau^{2})$$

(10)

Here, $\tau^{2}$ represents the between-study variance, capturing the variability in the true mean ratios ($\theta_{ratio, k}$) across studies. The parameters of these distributions were inferred with MCMC methods. We specified uninformative priors for each parameter, as proposed by J. P. Higgins et al. (25). The exponential and the lognormal distributions were visually compared to point estimates from individual studies with a cumulative distribution function (CDF). The analysis was performed using the *metafor* package version 3.0-2 (20) in R (version 4.3.0).

##### Subgroup analyses

Subgroup analyses were conducted to assess potential sources of variability in the mean ratios across various subgroups. The pooled estimates, representing the aggregated outcomes from each subgroup, were obtained using a Bayesian random-effects model. In this model, both the within-study mean and the random effect were assumed to be normally distributed.

---------------------------------------------------------------------------------------------------------------------

**Amendment #3: Omission of the sensitivity analysis**

Our original protocol outlined plans to conduct sensitivity analyses to investigate the sources of heterogeneity within our data. However, upon review, we found this step to be unnecessary because:

- Risks of bias were very similar across all examined studies, substantially reducing the potential insights that could be gained from a sensitivity analysis based on the risk of bias categories.
- The decision to employ a modified statistical method to incorporate non-detects into the analyses eliminated the need to evaluate the impact of excluding all sample-specific ratios with a non-detect in culture or qPCR.

---------------------------------------------------------------------------------------------------------------------

**Amendment #4: Omission of the qualitative synthesis**

Initially, our protocol included conducting a qualitative synthesis of the studies included in the review. However, we found that most studies lacked sufficient qualitative information for such an analysis during the data extraction process. This lack of information would limit the depth and accuracy of a qualitative analysis. Consequently, we decided to focus on quantitative data where more comprehensive information was available.

### S2 Mathematical proof

This section presents the proof for the transformation of the e.s.e on the log_10_ scale (Equation 8).

Since $y= \mathrm{LRV}\left( x \right)=-\log_{10} \left( x \right)$ is a continuous monotonic decreasing function, it follows that a confidence interval for $\mathrm{LRV}\left( \bar{r} \right)=-\log_{10} \left( \bar{r} \right)$ is:

| $\mathrm{LRV}\left( \bar{r} \right) \epsilon\left( -\log_{10} \left( e^{\left( \mu_{ratio}+\frac{\sigma_{ratio}^{2}}{2} \right)+t_{\frac{\alpha}{2}}\cdot e.s.e.\left( \hat{\ln\left( \bar{r} \right)} \right)} \right), -\log_{10} \left( e^{\left( \mu_{ratio}+\frac{\sigma_{ratio}^{2}}{2} \right)- t_{\frac{\alpha}{2}}\cdot e.s.e.\left( \hat{\ln\left( \bar{r} \right)} \right)} \right) \right)$ $= \mathrm{LRV}\left( \bar{r} \right) \epsilon\left( -\frac{1}{\ln\left( 10 \right)}\ln\left( e^{\left( \mu_{ratio}+\frac{\sigma_{ratio}^{2}}{2} \right)+t_{\frac{\alpha}{2}}\cdot e.s.e.\left( \hat{\ln\left( \bar{r} \right)} \right)} \right), -\frac{1}{\ln\left( 10 \right)}\ln\left( e^{\left( \mu_{ratio}+\frac{\sigma_{ratio}^{2}}{2} \right)- t_{\frac{\alpha}{2}}\cdot e.s.e.\left( \hat{\ln\left( \bar{r} \right)} \right)} \right) \right)$ $= \mathrm{LRV}\left( \bar{r} \right) \epsilon\left( -\frac{1}{\ln\left( 10 \right)} \left( \left( \mu_{ratio}+\frac{\sigma_{ratio}^{2}}{2} \right)+t_{\frac{\alpha}{2}}\cdot e.s.e\left( \hat{\ln\left( \bar{r} \right)} \right) \right), -\frac{1}{\ln\left( 10 \right)} \left( \left( \mu_{ratio}+\frac{\sigma_{ratio}^{2}}{2} \right)-t_{\frac{\alpha}{2}}\cdot e.s.e.\left( \hat{\ln\left( \bar{r} \right)} \right) \right) \right)$ $= \mathrm{LRV}\left( \bar{r} \right) \epsilon\left( \mathrm{LRV}\left( \bar{r} \right)+t_{\alpha/2}\cdot\mathrm{LRV}\left( e^{e.s.e.\left( \hat{\ln\left( \bar{r} \right)} \right)} \right) , \mathrm{LRV}\left( \bar{r} \right)-t_{\alpha/2}\cdot\mathrm{LRV}\left( e^{e.s.e\left( \hat{\ln\left( \bar{r} \right)} \right)} \right) \right)$ | (S.1) |
| --- | --- |

Note that the bounds of the confidence interval switch because $y=LRV(x)$ is a decreasing function.

It can be proved that $e.s.e.\left( \mathrm{LRV}\left( \bar{r} \right) \right)=\mathrm{LRV}\left( e^{e.s.e.\left( \hat{\ln\left( \bar{r} \right)} \right)} \right)$ as follows:

It is known that:

| $\mathrm{LRV}\left( \bar{r} \right)=-\log_{10} \left( e^{\left( \hat{\ln\left( \bar{r} \right)} \right)} \right)$ $= -\frac{1}{\ln\left( 10 \right)}\ln\left( e^{\left( \hat{\ln\left( \bar{r} \right)} \right)} \right)$ $=-\frac{1}{\ln\left( 10 \right)} \hat{\ln\left( \bar{r} \right)}$ $=c \cdot\hat{\ln\left( \bar{r} \right)}$ | (S.2) |
| --- | --- |

and from the rules of variance, we know that

$$\mathrm{Var}(\mathrm{cX})=c^{2} \cdot\mathrm{Var}(X)$$

and thus

$$\mathrm{sd}(\mathrm{cX})=\left| c \right| \mathrm{sd}(X)$$

Therefore

$$e.s.e\left( \mathrm{LRV}\left( \bar{r} \right) \right)=\mathrm{sd}\left( \mathrm{LRV}\left( \bar{r} \right) \right)$$

$$= \mathrm{sd}\left( -\frac{1}{\ln\left( 10 \right)} \hat{\ln\left( \bar{r} \right)} \right)$$

$$= \frac{1}{\ln\left( 10 \right)}\mathrm{sd}\left( \hat{\ln\left( \bar{r} \right)} \right)$$

$$= \frac{1}{\ln\left( 10 \right)}e.s.e\left( \hat{\ln\left( \bar{r} \right)} \right)$$

$$= \frac{1}{\ln\left( 10 \right)}\ln\left( e^{e.s.e\left( \hat{\ln\left( \bar{r} \right)} \right)} \right)$$

$$= \left| \mathrm{LRV}\left( e^{e.s.e\left( \hat{\ln\left( \bar{r} \right)} \right)} \right) \right|$$

**S3 Additional results**

This section provides additional figures and tables not included in the main paper.

**Table S1.** Comparison of Poisson gamma (PGA) distribution to the Poisson lognormal (PLN) distribution for predicting site-specific concentrations of *Legionella pneumophila* measured by qPCR in building drinking water systems. The marginal deviance information (mDIC) and its standard error (SE) are listed for each site. The lowest mDIC indicates the best model fit. A difference of 3.0 and more is considered significant. Bold mDIC values indicate best fit models.

| **Reference, site** | $\boldsymbol{n}$ | | |  | | **Parameter** | | | |  | **Deviance information criterion** | | | | |
| --- | --- | --- | --- | --- | --- | --- | --- | --- | --- | --- | --- | --- | --- | --- | --- |
|  | **Total** | **% of +ve qPCR** |  | | **PGA** | | | **PLN** | |  | | **PGA** | | **PLN** | |
|  |  |  |  | | $\hat{\boldsymbol{\alpha}}$ | | $\hat{\boldsymbol{\beta}}$ | $\hat{\boldsymbol{\mu}}$ | $\hat{\boldsymbol{\sigma}}$ |  | | **mDIC** | **SE** | **mDIC** | **SE** |
| Bonetta et al. (2018) Before inter. | 13 | 92 |  | | 0.9 | | 1.9E-5 | 10.1 | 1.7 |  | | - | - | **-** | - |
| Bonetta et al. (2018) After interv. | 51 | 45 |  | | 0.8 | | 3.4E-3 | 4.8 | 0.9 |  | | - | - | - | - |
| Grimard-Conea (2022) First draw | 62 | 100 |  | | 0.2 | | 5.8E-6 | 7.7 | 2.2 |  | | - | - | - | - |
| Joly et al. (2006), Grenoble | 31 | 100 |  | | 0.6 | | 6.0E-5 | 8.4 | 1.3 |  | | - | - | - | - |
| Lee et al. (2011), France DW1 | 36 | 67 |  | | 0.5 | | 1.9E-4 | 6.5 | 1.9 |  | | 377.8 | 0.4 | **367.7** | 0.4 |
| Lee et al. (2011), France DW2 | 33 | 91 |  | | 0.4 | | 9.3E-5 | 7.0 | 1.8 |  | | - | - | - | - |
| Lee et al. (2011), HPA SH2 | 12 | 83 |  | | 1.4 | | 1.4E-3 | 6.5 | 1.0 |  | | 105.6 | 0.1 | 107.3 | 0.1 |
| Lee et al. (2011), Italy Scre | 11 | 100 |  | | 1.4 | | 4.6E-4 | 7.7 | 0.9 |  | | 123.2 | 0.2 | 121.0 | 0.2 |
| Lee et al. (2011), Italy Ed 1 | 30 | 100 |  | | 4.8 | | 5.0E-3 | 6.7 | 0.4 |  | | 259.6 | 0.4 | **244.8** | 1.4 |
| Lee et al. (2011), Italy Pad 5 | 35 | 89 |  | | 0.4 | | 8.0E-5 | 7.3 | 1.8 |  | | 611.3 | 0.8 | **428.3** | 0.6 |
| Lee et al. (2011), Spain DW1 | 44 | 89 |  | | 0.5 | | 1.0E-4 | 7.6 | 1.5 |  | | - | - | **-** | - |
| Lee et al. (2011), Spain DW2 | 45 | 100 |  | | 2.0 | | 1.4E-3 | 7.0 | 0.6 |  | | 425.9 | 0.4 | **419.2** | 0.7 |
| Lee et al. (2011), Spain DW3 | 45 | 100 |  | | 0.9 | | 4.1E-5 | 9.5 | 1.2 |  | | 730.3 | 0.9 | **678.9** | 0.8 |
| Lee et al. (2011), Switz. 100 | 30 | 43 |  | | 0.4 | | 1.8E-4 | 6.1 | 1.4 |  | | - | - | **-** | - |
| Lee et al. (2011), Switz. 110 | 27 | 93 |  | | 0.6 | | 5.0E-5 | 8.3 | 1.7 |  | | 426.9 | 0.7 | **364.8** | 0.6 |
| Lee et al. (2011), Switz. 120 | 29 | 97 |  | | 0.4 | | 9.2E-7 | 11.3 | 2.5 |  | | - | - | **-** | - |
| Morio et al. (2008) | 98 | 55 |  | | 0.51 | | 3.1E-4 | 6.1 | 1.5 |  | | - | - | **-** | - |

**Table S2.** Comparison of Poisson gamma (PGA) distribution to the Poisson lognormal (PLN) distribution for predicting site-specific concentrations of *Legionella pneumophila* measured by culture in building drinking water systems. The marginal deviance information (mDIC) and its standard error (SE) are listed for each site. The lowest mDIC indicates the best model fit. A difference of 3.0 and more is considered significant. Bold mDIC values indicate best fit models.

| **Reference, site** | $\boldsymbol{n}$ | | |  | | **Parameter** | | | |  | **Deviance information criterion** | | | |
| --- | --- | --- | --- | --- | --- | --- | --- | --- | --- | --- | --- | --- | --- | --- |
|  | **Total** | **% of +ve culture** |  | | **PGA** | | | **PLN** | |  | **PGA** | | **PLN** | |
|  |  |  |  | | $\hat{\boldsymbol{\alpha}}$ | | $\hat{\boldsymbol{\beta}}$ | $\hat{\boldsymbol{\mu}}$ | $\hat{\boldsymbol{\sigma}}$ |  | **mDIC** | **SE** | **mDIC** | **SE** |
| Bonetta et al. (2018) Before inter. | 13 | 100 |  | | 0.5 | | 5.6E-5 | 8.1 | 2.2 |  | - | - | - | - |
| Bonetta et al. (2018) After interv. | 51 | 67 |  | | 0.1 | | 1.3E-3 | 1.2 | 2.8 |  | - | - | - | - |
| Grimard-Conea (2022) First draw | 62 | 69 |  | | 0.2 | | 9.0E-5 | 4.2 | 3.4 |  | 712.4 | 1.0 | **608.3** | 1.1 |
| Joly et al. (2006), Grenoble | 31 | 87 |  | | 0.5 | | 1.6E-4 | 6.6 | 1.9 |  | - | - | - | - |
| Lee et al. (2011), France DW1 | 36 | 47 |  | | 1.9 | | 3.6E-3 | 6.0 | 0.7 |  | 270.2 | 0.2 | **263.6** | 0.1 |
| Lee et al. (2011), France DW2 | 33 | 79 |  | | 0.6 | | 3.5E-4 | 6.4 | 1.1 |  | 748.3 | 2.0 | **558.1** | 1.2 |
| Lee et al. (2011), HPA SH2 | 12 | 50 |  | | 0.4 | | 2.4E-4 | 6.4 | 1.5 |  | 128.2 | 0.4 | **116.1** | 0.2 |
| Lee et al. (2011), Italy Scre | 11 | 64 |  | | 0.4 | | 1.1E-4 | 6.9 | 1.7 |  | 144.4 | 0.4 | **122.0** | 0.2 |
| Lee et al. (2011), Italy Ed 1 | 30 | 67 |  | | 2.3 | | 3.5E-3 | 6.3 | 0.7 |  | 238.7 | 0.1 | **233.5** | 0.1 |
| Lee et al. (2011), Italy Pad 5 | 35 | 69 |  | | 0.4 | | 8.0E-5 | 7.2 | 1.6 |  | 389.8 | 0.4 | 389.0 | 0.4 |
| Lee et al. (2011), Spain DW1 | 44 | 80 |  | | 0.8 | | 3.4E-4 | 7.0 | 1.2 |  | 570.3 | 1.3 | **457.5** | 0.5 |
| Lee et al. (2011), Spain DW2 | 45 | 58 |  | | 1.3 | | 1.6E-3 | 6.2 | 0.9 |  | 372.7 | 0.3 | **367.9** | 0.3 |
| Lee et al. (2011), Spain DW3 | 45 | 76 |  | | 0.5 | | 6.3E-5 | 7.7 | 1.8 |  | 560.5 | 0.6 | **555.2** | 0.6 |
| Lee et al. (2011), Switz. 100 | 30 | 20 |  | | 2.4 | | 8.0E-3 | 5.5 | 0.6 |  | 194.5 | 0.1 | **180.1** | 0.1 |
| Lee et al. (2011), Switz. 110 | 27 | 30 |  | | 2.0 | | 5.2E-3 | 5.6 | 0.7 |  | 186.3 | 0.1 | **177.5** | 0.1 |
| Lee et al. (2011), Switz. 120 | 29 | 38 |  | | 0.4 | | 1.9E-4 | 6.5 | 1.5 |  | 313.4 | 0.6 | **286.7** | 0.5 |
| Morio et al. (2008) | 98 | 31 |  | | 0.4 | | 1.0E-3 | 4.5 | 1.5 |  | 639.0 | 1.5 | **546.1** | 0.9 |

| **Log_10_ concentrations of *Legionella pneumophila* (CFU L^-1^)** | **A**. Lee et al. (2011) – Switz. 100 | **B.** Lee et al. (2011) – Switz. 110 | **C.** Lee et al. (2011) – Switz. 120 |
| --- | --- | --- | --- |
|  | 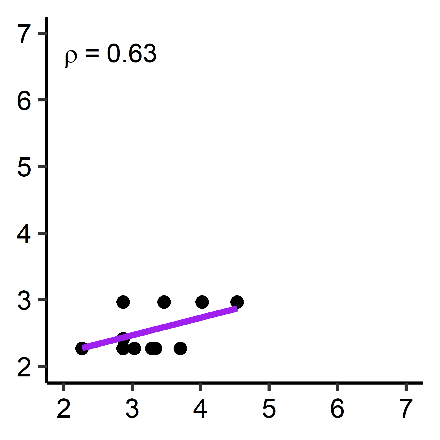 | 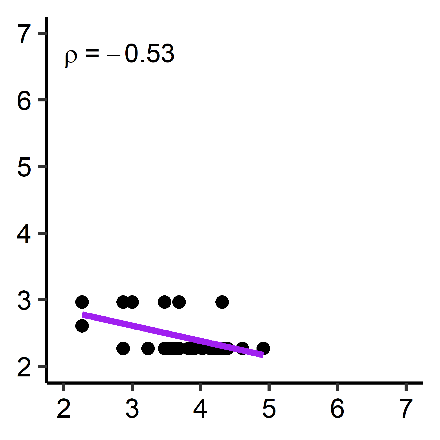 | 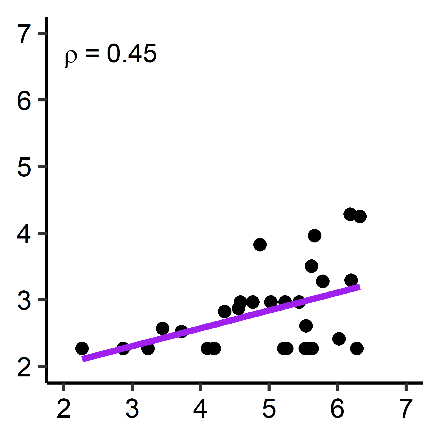 |
|  | **D.** Lee et al. (2011) – Spain DW1 | **E.** Lee et al. (2011) – Spain DW2 | **F.** Lee et al. (2011) – Spain DW3 |
|  | 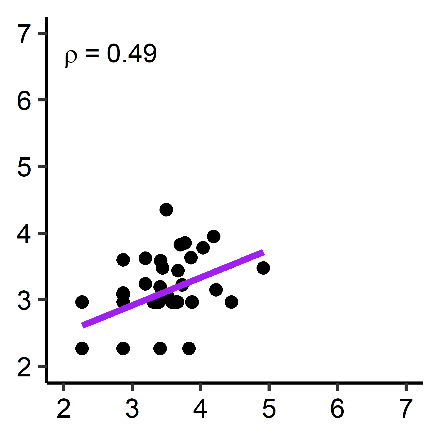 | 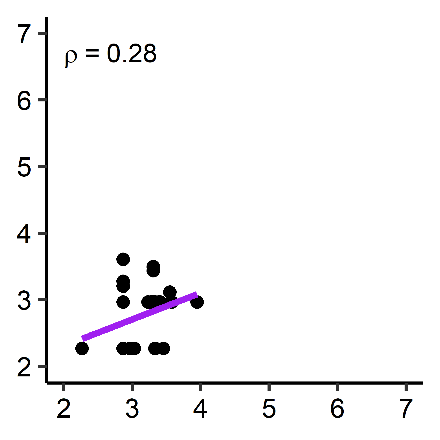 | 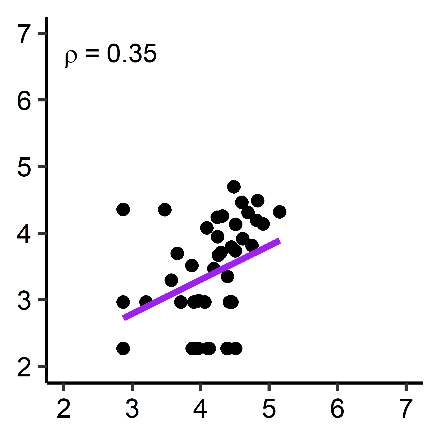 |
|  | **G.** Lee et al. (2011) – France DW1 | **H.** Lee et al. (2011) – France DW2 | **I.** Lee et al. (2011) – Italy Pad 5 |
|  | 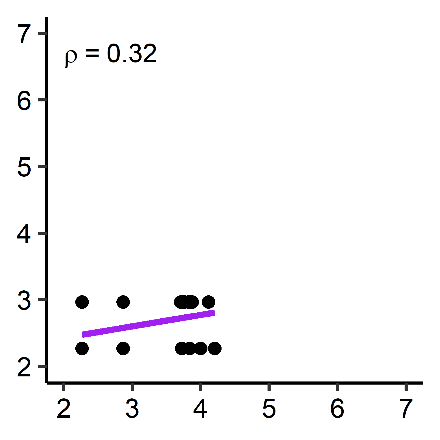 | 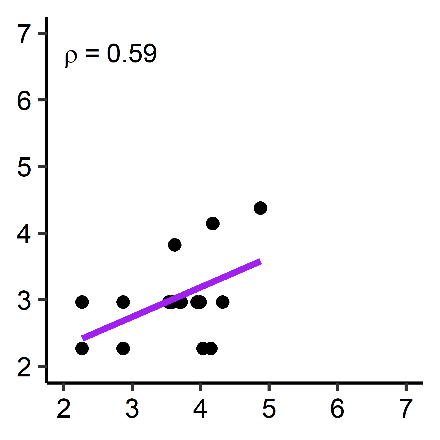 | 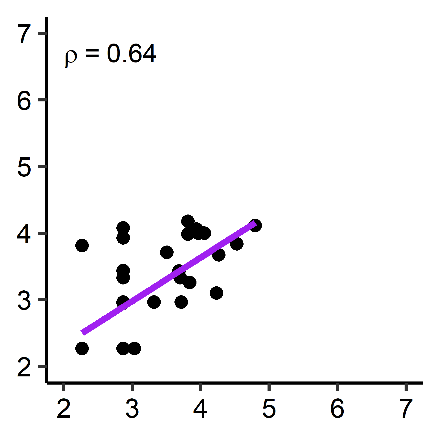 |
|  | **J.** Grimard-Conea et al. (2022) (a) | **K.** Joly et al. (2006) – Grenoble | **L.** Morio et al. (2008) |
|  | 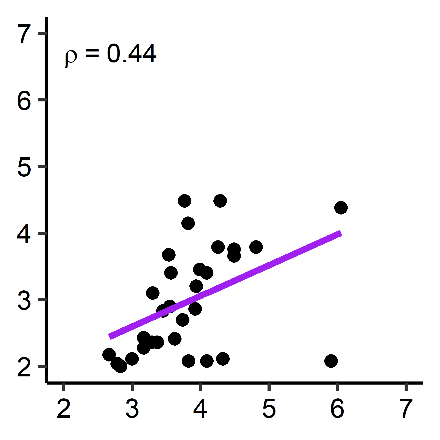 | 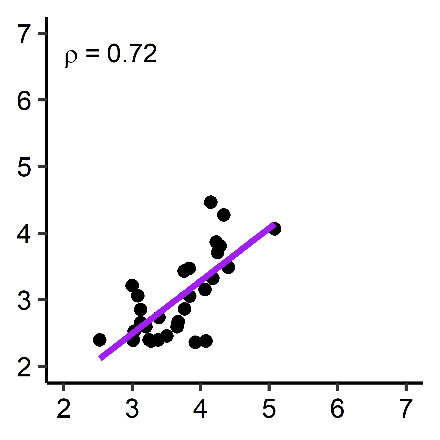 | 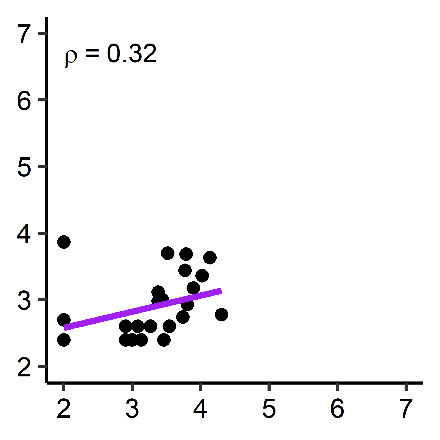 |
|  | **Log_10_ concentrations of *Legionella pneumophila* (GU L^-1^)** | | |

**Fig. S1.** Log-log regression model predicting culture concentrations from qPCR concentrations.

| **Author(s) and Year** | $\boldsymbol{n}$ $\boldsymbol{\mu}$ $\boldsymbol{\sigma}$ | **Log_10_ mean ratio [95% CI]** |
| --- | --- | --- |
| 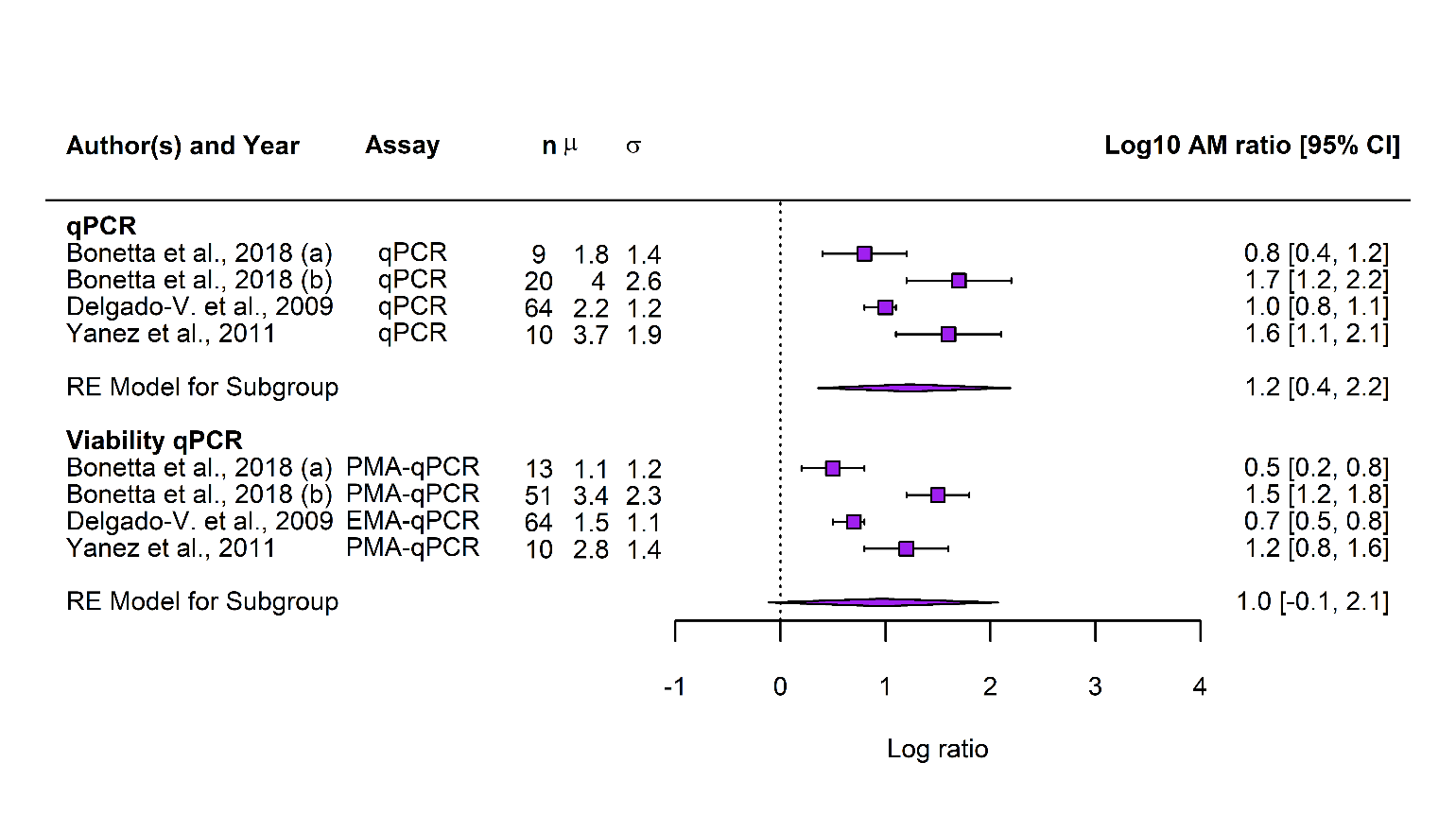 | | |
| **Log_10_ geometric mean qPCR:culture ratio** | | |

(a) No water treatment, (b) On-site water treatment with a neutral electrolyzed oxidizing water (NEOW) device

**Fig.S2.** Forest plots of qPCR:culture ratio and viability-qPCR:culture ratio for Legionella pneumophila in water samples from drinking water systems. Subgroup analyses were carried out for geometric mean ratios. Horizontal lines represent 95% confidence intervals on mean values. Pooled estimates were obtained using the random-effect model.

**Table S3.** Comparison of sample statistics and parameter values of lognormal distributions of the qPCR:culture ratio, qPCR concentrations and culture concentrations for measurement of *L. pneumophila* with standard qPCR and viability qPCR in paired water samples collected in potable water systems from reviewed studies.

| **Reference, site** | $\boldsymbol{n}$ | | |  | **Lognormal parameters** | | | | | | | **Pearson corr.**  **(**$\boldsymbol{\rho}$**)** | **qPCR:culture ratio** | |
| --- | --- | --- | --- | --- | --- | --- | --- | --- | --- | --- | --- | --- | --- | --- |
|  | **Total** | **% of +ve qPCR** | **% of +ve culture** |  | **qPCR** | | **Culture** | | **Ratio** | | |  | **Geometric mean** | **Arithmetic mean** |
|  |  |  |  |  | $\hat{\boldsymbol{\mu}}$ | $\hat{\boldsymbol{\sigma}}$ | $\hat{\boldsymbol{\mu}}$ | $\hat{\boldsymbol{\sigma}}$ | $\hat{\boldsymbol{\mu}}$ | $\hat{\boldsymbol{\sigma}}$ | ${\hat{\boldsymbol{\sigma}}}_{\boldsymbol{\rho}}$ |  |  |  |
| **qPCR** | | | | | | | | | | | | | | |
| Bonetta et al. (2018) (a) | 9 | 100 | 100 |  | 10.3 | 1.3 | 8.5 | 1.2 | 1.8 | 1.9 | 1.4 | 0.47 | 6 | 17 |
| Bonetta et al. (2018) (b) | 20 | 60 | 60 |  | 4.9 | 0.8 | 0.8 | 2.5 | 4.0 | 2.6 | 2.6 | 0.01 | 59 | 2100 |
| Delgado-V. et al. (2009) | 64 | 100 | 95 |  | 9.3 | 1.5 | 7.0 | 1.9 | 2.2 | 2.4 | 1.2 | 0.76 | 10 | 22 |
| Yanez et al. (2011) | 10 | 100 | 80 |  | 9.4 | 1.8 | 5.7 | 1.6 | 3.7 | 2.5 | 1.9 | 0.41 | 41 | 270 |
| **Viability qPCR** | | | | | | | | | | | | | | |
| Bonetta et al. (2018) (a) | 9 | 100 | 100 |  | 9.7 | 1.3 | 8.5 | 1.2 | 1.1 | 1.9 | 1.2 | 0.58 | 3 | 7 |
| Bonetta et al. (2018) (b) | 20 | 20 | 60 |  | 4.4 | 0.2 | 1.0 | 2.3 | 3.4 | 2.3 | 2.3 | $-$0.02 | 30 | 510 |
| Delgado-V. et al. (2009) | 64 | 95 | 95 |  | 8.5 | 1.6 | 7.0 | 1.9 | 1.5 | 2.5 | 1.1 | 0.83 | 4 | 10 |
| Yanez et al. (2011) | 10 | 100 | 80 |  | 7.6 | 1.0 | 4.8 | 1.3 | 2.8 | 1.6 | 1.4 | 0.26 | 16 | 48 |

**Table S4.** Selected quality control and assurance elements for qPCR analyses reported in reviewed studies.

|  | **Whole process** | |  | | **Nucleic acid extraction** | |  | **PCR detection** | | | |
| --- | --- | --- | --- | --- | --- | --- | --- | --- | --- | --- | --- |
| **Reference** | **Recovery control** |  | | **Recovery control** | | **Positive/negative control** |  | **Recovery control** | **Positive/negative control** | **Method for generating standard curve** | **Inhibition control** |
| *Site-specific data* | | | | | | | | | | | |
| Bonetta et al. (2018) | No |  | | No | | No/No |  | No | No/No | No | No |
| Grimard-C. et al. (2022) | No |  | | No | | No/No |  | No | No/No | Yes | Yes |
| Joly et al. (2006) | No |  | | No | | Yes/Yes |  | No | Yes/Yes | Yes | Yes |
| Lee et al. (2011)*^a^* | No |  | | Yes | | Yes/Yes |  | Yes | Yes/Yes | Yes | Yes |
| Morio et al. (2008) | No |  | | No | | No/No |  | No | No/Yes | Yes | Yes |
| *Pooled data* | | | | | | | | | | | |
| Behets et al. (2007) | Yes |  | | No | | Yes/Yes |  | No | Yes/Yes | Yes | Yes |
| Bonetta et al. (2010) | No |  | | No | | No/No |  | No | Yes/Yes | No | Yes |
| Boss et al. (2018) | No |  | | No | | Yes/No |  | No | Yes/Yes | Yes | No |
| Collins et al. (2015) | No |  | | Yes | | No/No |  | No | Yes/Yes | Yes | Yes |
| Collins et al. (2017) | No |  | | Yes | | No/No |  | No | Yes/Yes | Yes | Yes |
| Delgado-V. et al. (2009) | No |  | | No | | No/No |  | No | Yes/Yes | No | Yes |
| Fittipaldi et al. (2010) | No |  | | No | | No/No |  | No | No/No | No | No |
| Mapili et al. (2020) | No |  | | No | | No/No |  | No | No/No | No | Yes |
| Toplitsch et al. (2018) | No |  | | Yes | | No/No |  | No | Yes/Yes | Yes | Yes |
| Yanez et al. (2011) | No |  | | No | | No/No |  | No | Yes/Yes | No | Yes |

*^a^*Adhered to AFNOR NF-T90-471:2010

**Table S5.** Standard for cultivation of *Legionella pneumophila* reported in reviewed studies.

| **Reference** | **Standard** |
| --- | --- |
| *Site-specific data* | |
| Bonetta et al. (2018) | ISO 11731 |
| Grimard-C. et al. (2022)*^a^* | Not reported |
| Joly et al. (2006) | AFNOR NF T90-431 |
| Lee et al. (2011) | ISO 11731 |
| Morio et al. (2008) | AFNOR NF T90-431 |
| *Pooled data* | |
| Behets et al. (2007) | NEN 6265 |
| Bonetta et al. (2010) | ISO 11731 |
| Boss et al. (2018) | ISO 11731 |
| Collins et al. (2015) | ISO 11731 |
| Collins et al. (2017) | ISO 11731 |
| Delgado-V. et al. (2009) | AFNOR NF T90-431 |
| Fittipaldi et al. (2010) | ISO 11731 |
| Mapili et al. (2020)*^a^* | Not reported |
| Toplitsch et al. (2018) | ISO 11731 |
| Yanez et al. (2011) | ISO 11731 |

*^a^* The standard ASTM D8429-21 has been published in 2022 for the Legiolert^®^ test used in this study.
